# Cerebrospinal fluid haemoglobin as a monitoring biomarker for secondary brain injury after aneurysmal subarachnoid haemorrhage: prospective international multicentre validation study (HeMoVal)

**DOI:** 10.64898/2026.01.15.26343790

**Authors:** Kevin Akeret, Raphael M. Buzzi, Thomas Gentinetta, Moritz Saxenhofer, David Kronthaler, Elisa Colombo, Alexandra Grob, Bart Thomson, Nina Schwendinger, Amr Abdulazim, Joshua Haegler, Gwendoline Canzanella, Vincens Kälin, Linda Bättig, Lucas Moritz Wiggenhauser, Maria Wostrack, Carolin Albrecht, Matthias Gmeiner, Julia Shawarba, Daniel Couto, Sandra Wymann, Andreas Wassmer, Marlies Illi, Kathrin Bieri, Karl Roessler, Andreas Gruber, Bernhard Meyer, Constantin Roder, Isabel Charlotte Hostettler, Basil E. Grüter, Nima Etminan, Luca Regli, Emanuela Keller, Ulrike Held, Dominik J. Schaer, Michael Hugelshofer, the HeMoVal Research Group

## Abstract

**Objectives:** To validate whether cerebrospinal fluid oxyhaemoglobin (CSF-Hb), measured from external ventricular or lumbar drains, is associated with secondary brain injury (SAH-SBI) after aneurysmal subarachnoid haemorrhage (aSAH), and to assess its value as a real-time monitoring biomarker.

**Design:** Preregistered multicentre prospective observational cohort study.

**Setting:** Eight neurosurgical tertiary centres in Switzerland, Germany, and Austria, between August 2021 and June 2024.

**Participants:** 366 patients with aSAH (mean age 58 years; 65% women). Of these, 260 provided cerebrospinal fluid (CSF) samples via external ventricular drain (EVD; 2,467 samples, median 10 days per patient) and 66 via lumbar drain (LD; 379 samples, median 6 days).

**Interventions:** Daily CSF samples were collected via EVD or LD from day 1 to day 14 after haemorrhage; no therapeutic interventions were tested.

**Main outcome measures:** CSF-Hb and its metabolites were analysed post hoc in a blinded manner. The primary outcome was SAH-SBI, defined as a composite of angiographic vasospasm (aVSP), delayed cerebral ischaemia (DCI), and delayed ischaemic neurological deficits (DIND), assessed daily over 14 days. Secondary outcomes included temporal CSF-Hb profiles and associations with aneurysm location, haematoma volume, intraventricular haemorrhage, chronic hydrocephalus, and 3-month functional outcome.

**Results:** CSF-Hb showed a delayed peak pattern: concentrations were low after aSAH, rose to a maximum on day 10 (EVD-derived CSF-Hb median 11.3 µM, IQR 2.64 to 25.90), and then declined. Larger haematoma volume (p<0.001) and intraventricular haemorrhage (p<0.001) were associated with higher EVD-derived CSF-Hb. SAH-SBI occurred in 209/366 patients (57%). Daily EVD-derived CSF-Hb showed no association with SAH-SBI (p=0.25) and only poor prognostic potential of same-day SAH-SBI (area under the curve 0.59, 95% confidence interval 0.56–0.63), with substantial between-centre heterogeneity. The oxidised haemoglobin metabolite methaemoglobin was positively associated with SAH-SBI (p=0.023; odds ratio 1.18 per log[µM], 95% confidence interval 1.02–1.36). Acute-phase EVD-derived CSF-Hb correlated with chronic hydrocephalus (p=0.012) and poor 3-month functional outcome (p=0.008). Catheter-related infection rates were low (2.2%).

**Conclusions:** In this preregistered multicentre validation study, EVD-derived CSF-Hb did not perform as a robust real-time monitoring biomarker for SAH-SBI, showing limited same-day discrimination and substantial between-centre heterogeneity. These findings argue against clinical implementation of CSF-Hb point-measurement as a single-parameter biomarker. In contrast, CSF methaemoglobin remained consistently associated with SAH-SBI, supporting the mechanistic relevance of haemolysis-related pathways. Future work using the HeMoVal biobank will apply multi-marker, pathway-level analyses to define haemolysis-related biomarker signatures and provide a platform for robust external validation of future candidates.

**Study registration:** ClinicalTrials.gov NCT04998370; date of registration 10 August 2021.

**Summary Boxes:** *What is already known on this topic:*

- Preclinical animal models link cell-free haemoglobin in cerebrospinal fluid (CSF-Hb) to secondary brain injury after aneurysmal subarachnoid haemorrhage (SAH-SBI).
- A single-centre study reported strong associations between daily external ventricular drain (EVD) derived CSF-Hb levels and SAH-SBI, and suggested a strong predictive potential(area under the curve 0.89).
- CSF-Hb monitoring has therefore been proposed as a bedside biomarker, but it has not undergone multicentre validation.

*What this study adds:*

- In a preregistered multicentre cohort of 366 patients from eight neurosurgical centres, once-daily EVD-derived CSF-Hb measurements showed poor same-day discrimination for SAH-SBI (area under the curve 0.59) and substantial between-centre heterogeneity.
- In contrast, CSF methaemoglobin was consistently associated with SAH-SBI, and higher acute-phase CSF-Hb was related to chronic hydrocephalus and worse 3-month functional outcome.
- These findings argue against routine adoption of CSF-Hb point-measurements as bedside single-analyte, while supporting haemolysis-related pathways as mechanistic targets.

## Introduction

Aneurysmal subarachnoid haemorrhage (aSAH) frequently leads to subarachnoid haemorrhage-related secondary brain injury (SAH-SBI), which significantly worsens outcome in up to two-thirds of patients despite aneurysm securing and modern neurocritical care.[1,2] SAH-SBI typically arises between days 4 and 14 after haemorrhage and encompasses angiographic vasospasm (aVSP), delayed cerebral ischaemia (DCI), and delayed ischaemic neurological deficits (DIND).[3,4] Identifying patients at highest risk during this window remains challenging: clinical examination, transcranial Doppler ultrasound, and imaging-based assessments have limited accuracy and are inconsistently applied across centres.[5]

Cell-free oxyhaemoglobin in cerebrospinal fluid (CSF-Hb) is a key candidate mediator of SAH-SBI.[6–8] After aneurysm rupture, blood disperses through the subarachnoid and ventricular CSF space. Subsequent red blood cell lysis releases haemoglobin, which drives nitric oxide depletion, oxidative stress, lipid peroxidation, and downstream microvascular dysfunction.[6–8] In a previous single-centre study, daily CSF-Hb sampled from external ventricular drains (EVD) showed strong associations with SAH-SBI and high diagnostic accuracy (area under the curve [AUC] = 0.89), suggesting that CSF-Hb could serve as a bedside monitoring biomarker for imminent secondary brain injury.[8] However, biomarkers that perform well in single-centre discovery cohorts frequently show attenuated effect sizes when subjected to independent multicentre validation, particularly when pre-analytical handling and endpoint assessment vary between sites.[9,10]

The Hemoglobin Monitoring after Subarachnoid Hemorrhage Validation (HeMoVal) study was therefore designed as a preregistered, prospective multicentre cohort to test the monitoring performance of CSF-Hb for SAH-SBI during the 14-day high-risk period after aSAH.[11] In parallel, HeMoVal was structured as a biobanking platform: daily CSF sampling with standardised processing creates a longitudinal archive in which haemoglobin species, their oxidative metabolites, scavenger proteins, inflammatory mediators, and markers of neuroglial injury can be studied in combination. The present report addresses the predefined primary question of whether CSF-Hb can serve as a robust real-time monitoring biomarker for SAH-SBI, while providing the foundation for subsequent pathway-level biomarker analyses using the HeMoVal biobank.

## Methods

The study followed a preregistered protocol published open access before recruitment.[11] All analyses and visualisations adhered to a detailed statistical analysis plan (https://doi.org/10.5281/zenodo.5556317). Reporting follows STROBE recommendations for observational studies.[12] Analyses beyond the prespecified plan are explicitly labelled as additional.

### Study design

This was a prospective, preregistered, international multicentre observational cohort study (ClinicalTrials.gov NCT04998370) conducted at eight academic neurosurgical tertiary care centres in Switzerland, Germany, and Austria.

### Ethics

The study was approved by the local research ethics committee at each participating centre (details in the supplementary material) and conducted in accordance with the Declaration of Helsinki and International Council for Harmonisation Good Clinical Practice guidelines (ICH-GCP E6[R2]). Written and oral study information was provided to patients or their legal representatives, and written informed consent was obtained. If patients initially unable to consent later declined participation, stored samples were destroyed and associated data anonymised and excluded. Data collected before withdrawal from previously consenting patients were retained in the analysis.

### Biobanking

In addition to the primary monitoring objective, the protocol prospectively mandated CSF biobanking for future mechanistic biomarker studies. Aliquots not required for the present CSF-Hb and metabolite measurements were stored centrally under standardised conditions as part of the HeMoVal biobank.

### Population

Adults (≥18 years) with radiologically confirmed subarachnoid haemorrhage were screened within 24 hours of admission. Exclusion criteria were non-aneurysmal SAH (e.g. traumatic or perimesencephalic), concurrent enrolment in an interventional or other CSF sampling study, or previous enrolment in HeMoVal.

### Objectives

The primary objective was to validate the association between morning EVD-derived CSF-Hb concentrations and SAH-SBI (assessed each afternoon over the preceding 24 hours) during the first 14 days after haemorrhage. Secondary objectives were to characterise temporal CSF-Hb profiles (EVD and LD), examine associations with aneurysm location, haematoma volume, SAH-SBI components (aVSP, DCI, DIND), chronic hydrocephalus, and 3-month functional outcome (GOSE, mRS), and to evaluate discrimination for SAH-SBI, aVSP, DCI, and DIND.

### Procedures and outcomes

CSF samples (2 ml) were collected once daily from day 1 to day 14 after haemorrhage via EVD or LD, when a CSF diversion device was present and sampling was not contraindicated. Samples were centrifuged immediately (1500 g, 15 minutes), the supernatant stored at -80°C, and batches shipped to the Research Biobanking Service Center, University Hospital Zurich, Switzerland. After completion of clinical follow-up, samples were transferred to CSL Behring, Berne, for central, blinded CSF-Hb measurements. Oxyhaemoglobin, methaemoglobin, bilirubin, and biliverdin were quantified using UV/Vis spectrophotometry (Lunatic, Unchained Labs; 230–750 nm) with spectral deconvolution as previously described.[6,8]

The primary endpoint, SAH-SBI, was assessed daily over 14 days and was defined as the composite of:

aVSP: arterial narrowing on CT angiography, MR angiography, or digital subtraction angiography;

DCI: a new ischaemic lesion or perfusion deficit;

DIND: a new focal neurological deficit or ≥2-point decrease in Glasgow Coma Scale lasting ≥2 hours.

Endpoints were classified as non-assessable if clinical or imaging data were insufficient. For the composite outcome, non-assessability was assigned if none of the component endpoints could be assessed.

Secondary endpoints included induced hypertension, spasmolysis, surgical decompression, and safety outcomes (CSF infection, revision surgery). Chronic hydrocephalus and functional outcomes (GOSE, mRS) were assessed at 3-month follow-up.

Endpoint assessors and treating teams had no access to CSF-Hb or metabolite concentrations, which were measured after completion of follow-up.

Baseline data included demographics, medical history, clinical status and scores, radiological features, and aneurysm treatment. Haematoma volume on baseline CT was quantified centrally using a published algorithm, blinded to clinical outcomes and CSF-Hb results.[13]

### Data management and oversight

Data management and monitoring were coordinated by the Department of Clinical Research, University of Bern, Switzerland. Electronic case report forms were implemented in REDCap and stored on a secure Linux-based MySQL server. Central and on-site monitoring and regular reviews of electronic records ensured data quality. CSF samples were barcode-labelled and linked to study identifiers in the database. Baseline CT scans were uploaded to a secure, HIPAA-compliant platform (Augsafe, Augmedit BV, Naarden, Netherlands) for central imaging analysis.

### Sample size

The primary objective focused on patients with EVDs; secondary objectives also included those with LDs. Sample size calculations assumed an anticipated effect size of OR 0.1 / µM CSF-Hb, 90% power, and between-site variability modelled in a generalised linear mixed model, as described.[11] This yielded a minimum of six sites and 30 EVD patients per site.[11] Allowing for a 15% dropout rate and variable recruitment, the final target was eight sites and 250 EVD patients.[11]

### Statistical analysis

All analyses were performed in R (version 4.5.2)[14] according to the prespecified statistical analysis plan.[11] Statistical code was developed and cross-checked by two authors using mock data (KA, RMB), with independent review by a senior statistician (UH). A second statistician independently reproduced the primary analyses (DK). The final analysis was conducted after database lock and data cleaning by the central data monitor. Additional exploratory analyses are labelled accordingly.

Three analysis cohorts were defined: the EVD cohort (≥1 EVD CSF sample), the LD cohort (≥1 LD CSF sample), and the full cohort (all enrolled patients). The null hypothesis was that there is no association between EVD-derived CSF-Hb concentration and SAH-SBI during the first 14 days after haemorrhage in the EVD cohort.

The primary analysis used a generalised additive model (GAM) to estimate the association between same-day SAH-SBI and EVD-derived CSF-Hb, with day after haemorrhage modelled as a continuous covariate (4-knot spline) and repeated measures nested within patients and sites via random intercepts. This modelling strategy was chosen a priori to emulate a potential bedside use case, focusing on within-patient, same-day associations while accounting for site-level differences.[11] Results are reported as linear terms, 95% confidence intervals, and p-values, and for smoothing splines the estimated functions with 95% confidence bands and p-values. For SAH-SBI, the AUC was calculated to quantify discriminative performance by integrating sensitivity versus 1−specificity across all possible CSF-Hb concentration thresholds.[11]

Secondary and safety analyses had no predefined significance thresholds, followed our preregistered protocol and are reported descriptively.[11] For associations between CSF-Hb and aneurysm location, baseline haematoma volume, intraventricular haemorrhage, and day after haemorrhage we used a generalised additive model with a 4-knot spline for time and random intercepts for patients and sites.

In an additional exploratory analysis, we fitted site-specific generalised additive models, including a linear term for log-transformed CSF-Hb, a penalised spline for time, and random intercepts for patients, omitting the site-level random effect to enable centre-specific estimates.

### Data availability

Anonymized participant data (https://doi.org/10.5281/zenodo.6609706) and the full analysis code (https://doi.org/10.5281/zenodo.6620697) are shared via a public repository. Participating centres are masked to preserve institutional confidentiality. Additionally, free-text comments containing potentially identifiable information (for example, explanations for omitted sampling) were removed to preserve anonymity.

### Patient and public involvement

Patients or members of the public were not involved in the design, conduct, reporting, or dissemination of this research.

## Results

### Patient characteristics and management

Between August 2021 and June 2024, 474 patients were screened and 108 were excluded (46 ineligible, 45 declined, 17 other reasons). The final cohort comprised 366 patients, including 260 with at least one EVD sample and 66 with at least one LD sample (figure 1). Demographic and clinical characteristics were comparable between cohorts, apart from intraventricular haemorrhage, which was more frequent in the EVD group (66.9% vs 45.5%), consistent with clinical indications for ventriculostomy (table 1, supplementary table 1). Mean age of the full cohort was 58.0 years (SD 13.2), and 35% of patients were male. Clinical severity at presentation was WFNS grade 1 in 33.9%, grade 2 in 23.2%, grade 3 in 3.6%, grade 4 in 20.2%, and grade 5 in 19.1%. Aneurysms were treated mainly by endovascular coiling (55.7%) or surgical clipping (36.6%). Protocol adherence was high, with only minor deviations evenly distributed across sites, and no withdrawals due to protocol violations.

**Figure 1.**
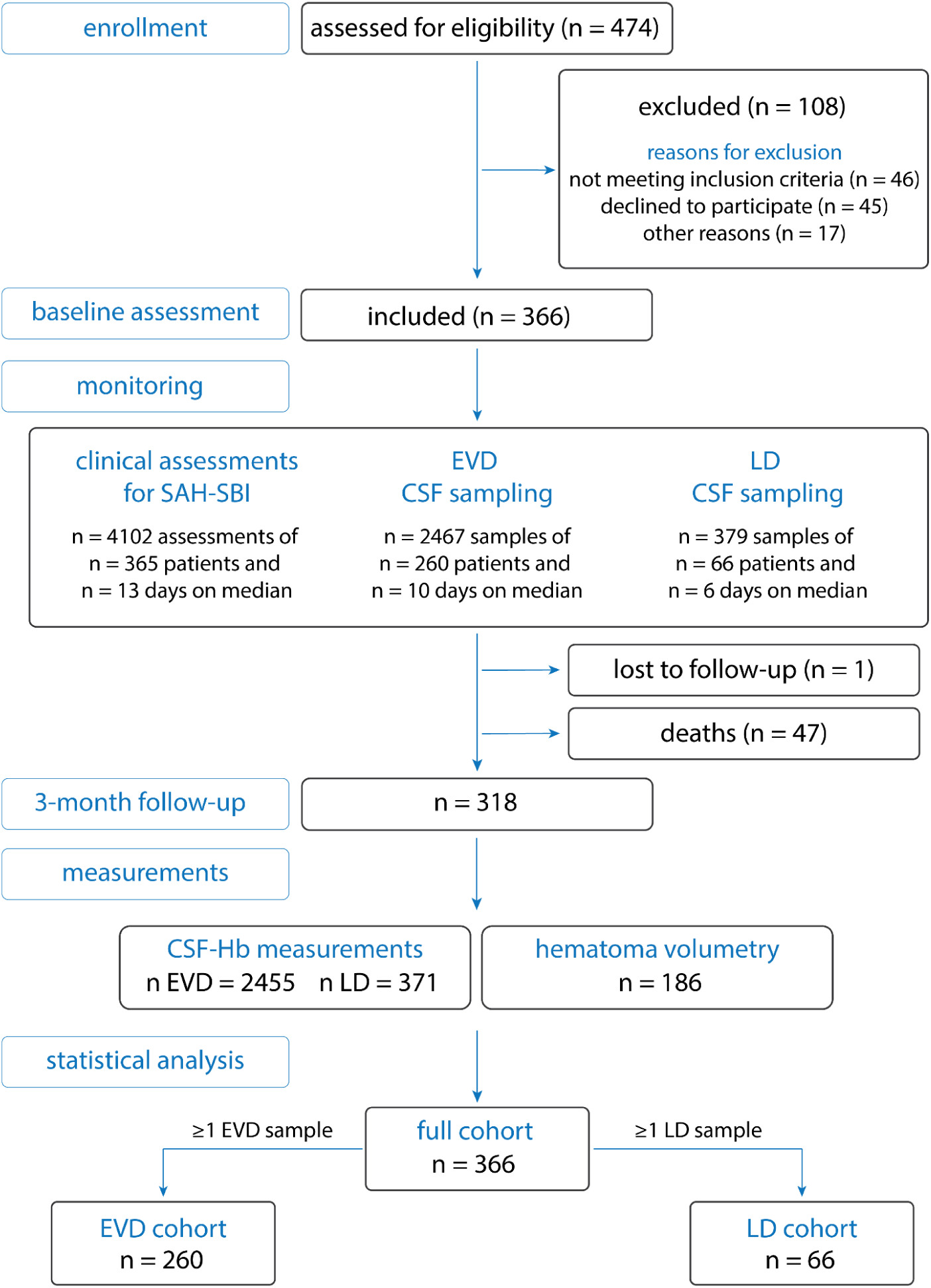
Study flowchart. Flow of patients through enrollment, baseline assessment, 14-day post-SAH monitoring (*daily CSF sampling in the morning, if an EVD or LD was in place, and clinical assessment in the afternoon*), and 3-month follow-up. The diagram also shows post hoc CSF analyses, haematoma volumetry, and definition of the three statistical analysis cohorts. *Abbreviations: aSAH, aneurysmal subarachnoid haemorrhage; CSF, cerebrospinal fluid; EVD, external ventricular drain; LD, lumbar drain; SAH-SBI, subarachnoid haemorrhage–related secondary brain injury*.

**Table 1.** Baseline characteristics of the study cohort. Baseline clinical and radiological characteristics of all enrolled patients. Characteristics of individual centre cohorts are shown in supplementary table 1. Values are presented as number (percentage) or mean (standard deviation). *Abbreviations: ACA, anterior cerebral artery; ACOM, Anterior communicating artery; EVD, external ventricular drain; LD, lumbar drain; MCA, middle cerebral artery; PCOM, posterior communicating artery; PICA, posterior inferior cerebellar artery; WFNS, World Federation of Neurosurgical Societies*. ^1^distal to the anterior communicating artery (e.g., pericallosal artery, callosomarginal artery); ^2^ophthalmic or superior hypophyseal artery; ^3^anterior choroidal artery, anterior inferior cerebellar artery, basilar artery aneurysm not involving the tip (e.g., pontine branches), non-paraophthalmic internal carotid artery (e.g., cavernous blisters), posterior cerebral artery, superior cerebellar artery, vertebral artery; ^4^maximal diameter in mm; ^5^a hematoma bursting into the ventricular system was considered an intraventricular haemorrhage, while a pure retrograde redistribution of blood into the ventricular system was not; ^6^in cm^3^ based on automated segmentation and volumetry.

|  |  | Full cohort | EVD cohort | LD cohort |
| --- | --- | --- | --- | --- |
| **n** |  | 366 | 260 | 66 |
| **Age** (mean, SD) |  | 58.0 (13.2) | 59.4 (13.3) | 57.5 (11.7) |
| **Male sex** (n, %) |  | 128 (35.0%) | 82 (31.5%) | 23 (34.8%) |
| **WFNS grade** (n, %) | *I* | 124 (33.9%) | 51 (19.6%) | 23 (34.8%) |
|  | *II* | 85 (23.2%) | 66 (25.4%) | 20 (30.3%) |
|  | *III* | 13 (3.6%) | 10 (3.8%) | 4 (6.1%) |
|  | *IV* | 74 (20.2%) | 65 (25.0%) | 12 (18.2%) |
|  | *V* | 70 (19.1%) | 68 (26.2%) | 7 (10.6%) |
| **Aneurysm location** (n, %) | *ACOM* | 130 (35.5%) | 92 (35.4%) | 27 (40.9%) |
|  | *Basilar tip* | 24 (6.6%) | 19 (7.3%) | 8 (12.1%) |
|  | *Distal ACA*<sup>1</sup> | 21 (5.7%) | 14 (5.4%) | 5 (7.6%) |
|  | *MCA* | 74 (20.2%) | 43 (16.5%) | 14 (21.2%) |
|  | *Paraophthalmic*<sup>2</sup> | 10 (2.7%) | 6 (2.3%) | 1 (1.5%) |
|  | *PCOM* | 41 (11.2%) | 30 (11.5%) | 5 (7.6%) |
|  | *PICA* | 18 (4.9%) | 18 (6.9%) | 2 (3.0%) |
|  | *Others*<sup>3</sup> | 48 (13.1%) | 38 (14.6%) | 4 (6.1%) |
| **Aneurysm size**<sup>4</sup> (mean, SD) |  | 7.1 (6.8) | 6.9 (6.1) | 6.3 (3.3) |
| **Intracerebral hemorrhage** (n, %) |  | 127 (34.7%) | 103 (39.6%) | 21 (31.8%) |
| **Intraventricular haemorrhage**<sup>5</sup> (n, %) |  | 197 (53.8%) | 174 (66.9%) | 30 (45.5%) |
| **Modified Fisher grade** (n, %) | *1* | 59 (16.1%) | 18 (6.9%) | 11 (16.7%) |
|  | *2* | 29 (7.9%) | 23 (8.8%) | 5 (7.6%) |
|  | *3* | 110 (30.1%) | 68 (26.2%) | 25 (37.9%) |
|  | *4* | 168 (45.9%) | 151 (58.1%) | 25 (37.9%) |
| **Hematoma volume**<sup>6</sup> (mean, SD) |  | 60.0 (54.5) | 60.0 (54.5) | 73.4 (96.4) |
| **Aneurysm therapy** (n, %) | *Bypass* | 3 (0.8%) | 2 (0.8%) | 0 (0%) |
|  | *Clipping* | 134 (36.6%) | 85 (32.7%) | 22 (33.3%) |
|  | *Coiling* | 204 (55.7%) | 152 (58.5%) | 36 (54.5%) |
|  | *Flow diverter* | 16 (4.4%) | 13 (5.0%) | 4 (6.1%) |
|  | *Others* | 7 (1.9%) | 6 (2.3%) | 4 (6.1%) |
|  | *None* | 2 (0.5%) | 2 (0.8%) | 0 (0%) |

### Time course of CSF-Hb after aSAH

In total, 2,467 EVD samples (median 10 days per patient) and 379 LD samples (median 6 days) were analysed (figure 1). EVD-derived CSF-Hb concentrations were initially low, increased to a peak on day 10 after haemorrhage (median 11.3 µM, IQR 2.64 to 25.90), and subsequently declined (figure 2A; supplementary table 2). LD-derived CSF-Hb showed a similar delayed peak pattern without significant differences compared with EVD-derived levels (figure 2B; supplementary table 2). Considerable between-centre variability in CSF-Hb concentrations was observed (supplementary figure 1; supplementary table 3).

**Figure 2.**
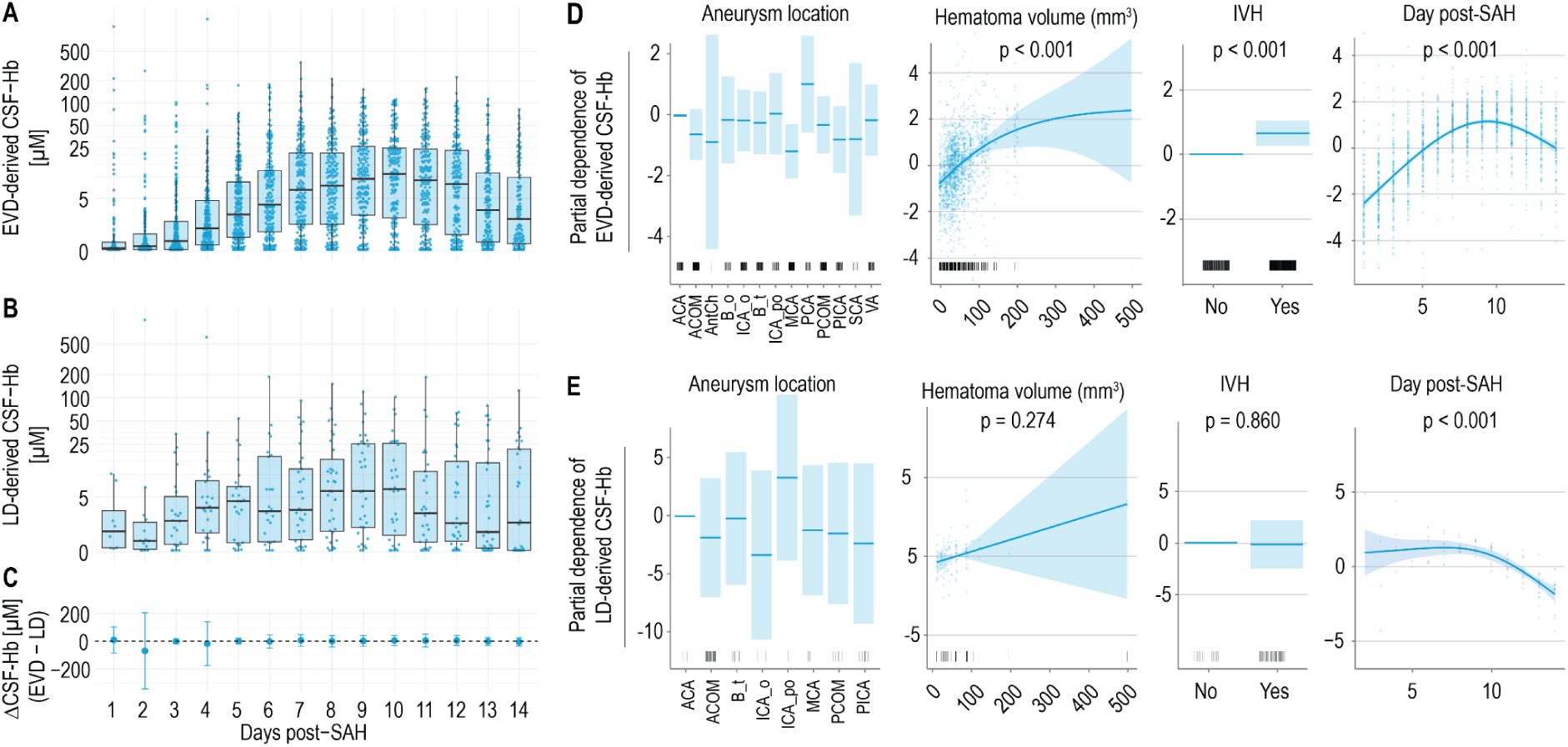
Temporal profiles and determinants of EVD- and LD-derived CSF-Hb after aSAH. **A.** Time course of EVD-derived CSF-Hb concentrations from day 1 to day 14 after aneurysmal subarachnoid haemorrhage (aSAH). **B.** Time course of LD-derived CSF-Hb concentrations over the same period. **C.** Daily differences between EVD- and LD-derived CSF-Hb concentrations after haemorrhage (mean difference ± SD). **D.** Generalised additive model (GAM) showing the dependence of EVD-derived CSF-Hb on aneurysm location, baseline haematoma volume, intraventricular haemorrhage (IVH), and day post-SAH. **E.** Corresponding GAM for LD-derived CSF-Hb. *Abbreviations: aSAH, aneurysmal subarachnoid haemorrhage; CSF-Hb, cerebrospinal fluid oxyhaemoglobin; EVD, external ventricular drain; LD, lumbar drain; IVH, intraventricular haemorrhage; GAM, generalised additive model*.

### Determinants of CSF-Hb concentrations

In the EVD cohort, CSF-Hb was significantly associated with baseline haematoma volume (p<0.001, figure 2D). A similar effect was observed in the LD cohort (figure 2E), although with lower evidence, likely due to limited sample size (p=0.274). Intraventricular haemorrhage was associated with higher EVD-derived CSF-Hb (effect size 0.66 log units, corresponding to ≈1.94-fold higher oxyhaemoglobin; 95% CI 1.30 to 2.87; p=0.001, figure 2D), with no effect in LD samples (effect size -0.21 log units, corresponding to ≈0.818-fold lower oxyhaemoglobin; 95% CI 0.08 to 8.02; p=0.86, figure 2E). EVD-derived CSF-Hb showed a delayed, parabolic relationship to post-haemorrhage day with a peak at day 10 (figure 2D), whereas LD-derived CSF-Hb remained constant until day 10 and declined thereafter (figure 2E). Aneurysm location was not associated with CSF-Hb, except for a negative association between ruptured middle cerebral artery aneurysms and EVD-derived CSF-Hb (p=0.008, figure 2D). No significant location effects were detected in LD samples (figure 2E).

### CSF-Hb and secondary brain injury

SAH-SBI occurred in 209/366 patients (57.1%) at least once, angiographic vasospasm in 169 (46.2%), DCI in 83 (22.7%), and DIND in 102 (27.9%) (table 2; supplementary table 4). Therapeutic interventions included induced hypertension (42.3%), spasmolysis (20.5%), and surgical decompression (11.7%), with similar frequencies in EVD and LD cohorts (table 2; supplementary table 4).

**Table 2.** SAH-SBI related outcomes and interventions. Incidence of subarachnoid haemorrhage–related secondary brain injury (SAH-SBI; composite of angiographic vasospasm [aVSP], delayed cerebral ischaemia [DCI], and delayed ischemic neurological deficits [DIND]), its individual components (aVSP, DCI, DIND), and related interventions (triple-H therapy: induced hypertension and/or hypervolaemia and/or haemodilution; spasmolysis; surgical decompression by hemicraniectomy or suboccipital craniectomy) in the full cohort and in the external ventricular drain (EVD) and lumbar drain (LD) subcohorts. Incidence is reported per patient and per assessment day (p, present; a, absent; m, missing).

| Outcome / Measure | Incidence |  |  |  |  |  |  |  |  |  |  |  |
| --- | --- | --- | --- | --- | --- | --- | --- | --- | --- | --- | --- | --- |
|  | Full cohort |  |  |  | EVD cohort |  |  |  | LD cohort |  |  |  |
|  | Per patient | Per day |  |  | Per patient | Per day |  |  | Per patient | Per day |  |  |
|  |  | a | p | m |  | a | p | m |  | a | p | m |
| SAH-SBI | 209<br>(57.1) | 3568<br>(70.8) | 534<br>(10.6) | 935<br>(18.6) | 154<br>(59.2) | 2358<br>(66.3) | 416<br>(11.7) | 780<br>(21.9) | 35<br>(53.0) | 704<br>(76.2) | 102<br>(11.0) | 118<br>(12.8) |
| aVSP | 169<br>(46.2) | 761<br>(15.1) | 367<br>(7.3) | 3909<br>(77.6) | 124<br>(47.7) | 612<br>(17.2) | 280<br>(7.9) | 2662<br>(74.9) | 30<br>(45.5) | 102<br>(11.0) | 64<br>(6.9) | 758<br>(82.0) |
| DCI | 83<br>(22.7) | 1105<br>(21.9) | 142<br>(2.8) | 3790<br>(75.2) | 70<br>(26.9) | 868<br>(24.4) | 124<br>(3.5) | 2562<br>(72.1) | 10<br>(15.2) | 173<br>(18.7) | 17<br>(1.8) | 734<br>(79.4) |
| DIND | 102<br>(27.9) | 3355<br>(66.6) | 228<br>(4.5) | 1454<br>(28.9) | 74<br>(28.5) | 2117<br>(59.6) | 171<br>(4.8) | 1266<br>(35.6) | 22<br>(33.3) | 702<br>(76.0) | 58<br>(6.3) | 164<br>(17.7) |
| Triple H therapy | 155<br>(42.3) | 3849<br>(76.4) | 1011<br>(20.1) | 177<br>(3.5) | 119<br>(45.8) | 2646<br>(74.5) | 839<br>(23.6) | 69<br>(1.9) | 25<br>(37.9) | 760<br>(82.3) | 142<br>(15.4) | 22<br>(2.4) |
| Spasmo-lysis | 75<br>(20.5) | 4677<br>(92.9) | 183<br>(3.6) | 177<br>(3.5) | 59<br>(22.7) | 3339<br>(94.0) | 146<br>(4.1) | 69<br>(1.9) | 12<br>(18.2) | 877<br>(94.9) | 25<br>(2.7) | 22<br>(2.4) |
| Surgical decompression | 43<br>(11.7) | 4815<br>(95.6) | 45<br>(0.9) | 177<br>(3.5) | 36<br>(13.8) | 3447<br>(97.0) | 38<br>(1.1) | 69<br>(1.9) | 5<br>(7.6) | 897<br>(97.1) | 5 (0.5) | 22<br>(2.4) |

Across 4,102 daily assessments (median 13 days per patient), we recorded 534 SAH-SBI events (10.6%), 367 aVSP events (7.3%), 142 DCI events (2.8%), and 228 DIND events (4.5%) (table 2). Event rates were low during the first three days, peaked around day 10, and declined thereafter (figure 3A). Non-assessable entries were most frequent for aVSP and DCI, which require imaging, whereas DIND had fewer non-assessable days, primarily when neurological examination was precluded by sedation (figure 3A).

**Figure 3.**
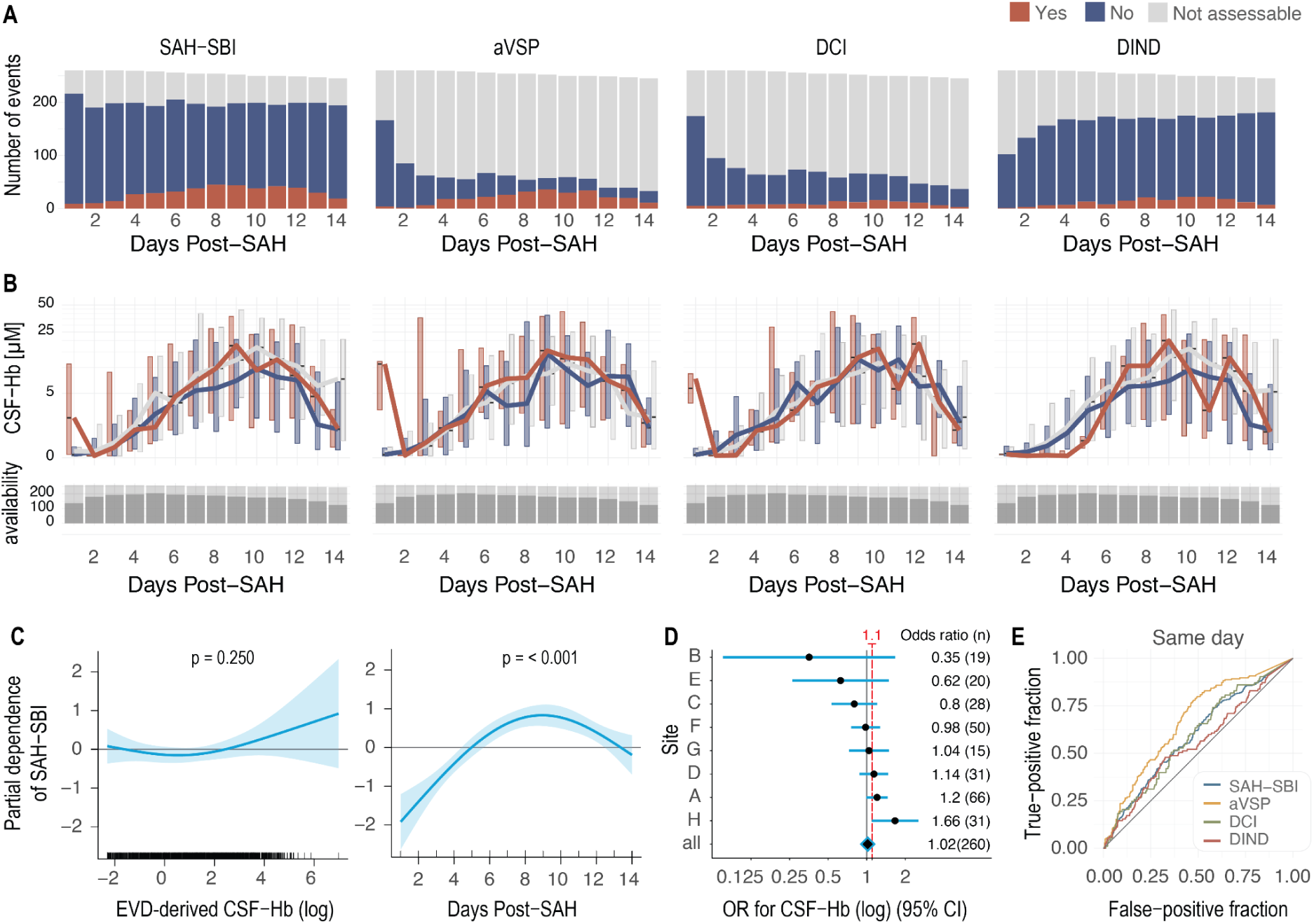
Association of EVD-derived CSF-Hb with secondary brain injury. **A.** Daily incidence of SAH-SBI and its components (aVSP, DCI, DIND) from day 1 to 14 after aSAH. **B.** Temporal trajectories of EVD-derived cerebrospinal fluid haemoglobin (CSF-Hb) stratified by event occurrence (top). Availability of CSF samples (bottom). **C.** Partial-dependence plots from generalised additive models (GAMs) showing no significant association between CSF-Hb and SAH-SBI (p=0.25), whereas post-haemorrhage day remained a strong covariate (p<0.001). **D.** Site-specific odds ratios for CSF-Hb and SAH-SBI, demonstrating pronounced heterogeneity across centres with 1.1 highlighted as the prespecified effect size. **E.** Receiver operating characteristic curves for same-day discrimination of CSF-Hb for SAH-SBI (AUC_SAH-SBI_ 0.59 [95% CI 0.56–0.63|, AUC_aVSP_ 0.60 [95% CI 0.56–0.64], AUC_DCI_ 0.56 [95% CI 0.51–0.61], and AUC_DIND_ 0.58 [95% CI 0.53–0.63]). *Abbreviations: aVSP, angiographic vasospasm; AUC, area under the curve; CSF-Hb, cerebrospinal fluid oxyhaemoglobin; DCI, delayed cerebral ischaemia; DIND, delayed ischemic neurological deficit; EVD, external ventricular drain; SAH-SBI, subarachnoid haemorrhage–related secondary brain injury*.

Descriptively, EVD-derived CSF-Hb tended to be higher in patients with SAH-SBI, aVSP, DCI, or DIND, compared to patients without these outcomes. CSF sampling coverage per day post-SAH was high throughout the observation window (figure 3B).

The generalised additive model for the EVD cohort did not suggest an association between EVD-derived CSF-Hb and SAH-SBI (p=0.25; figure 3C), whereas day after haemorrhage remained strongly associated with SAH-SBI (p<0.001). Similarly, no associations were observed between EVD-derived CSF-Hb and the individual components aVSP (p=0.37; supplementary figure 2A), DCI (p=0.64; supplementary figure 2B), or DIND (p=0.65; supplementary figure 2C).

The additional exploratory analysis to estimate centre-specific estimates revealed marked heterogeneity in the association between EVD-derived CSF-Hb and SAH-SBI (figure 3D). An effect size consistent with the anticipated odds ratio of about 1.1 per log(µM) increase in oxyhaemoglobin, based on the pilot cohort,[8] was seen at three sites, whereas centres with smaller numbers of patients showed attenuated or even inverse effects. The overall model yielded an odds ratio of 1.02 per log(µM) increase in oxyhaemoglobin (p=0.37).

Morning EVD-derived CSF-Hb showed poor discrimination for same-day SAH-SBI (AUC 0.59, 95% CI 0.56 to 0.63; figure 3E) and for same-day events during the high-risk phase from days 4 to 14 (AUC 0.55, 95% CI 0.52 to 0.59; supplementary figure 2D).

Results in the LD cohort were similar, with no significant associations (supplementary figure 3).

### CSF-Hb, chronic hydrocephalus, and 3-month outcome

At 3-months, one patient was lost to follow-up, 47 died, and 318 completed assessment (figure 1). Patients with chronic hydrocephalus had higher acute-phase CSF-Hb in both the EVD cohort (figure 4A; supplementary table 5) and LD cohort (supplementary figure 4A; supplementary table 5). Acute-phase CSF-Hb was also higher in patients with poor functional outcomes (GOSE 1-4 or mRS 4-6) compared with those with favourable outcomes (figure 4B-C; supplementary figure 4B-C; supplementary table 5).

**Figure 4.**
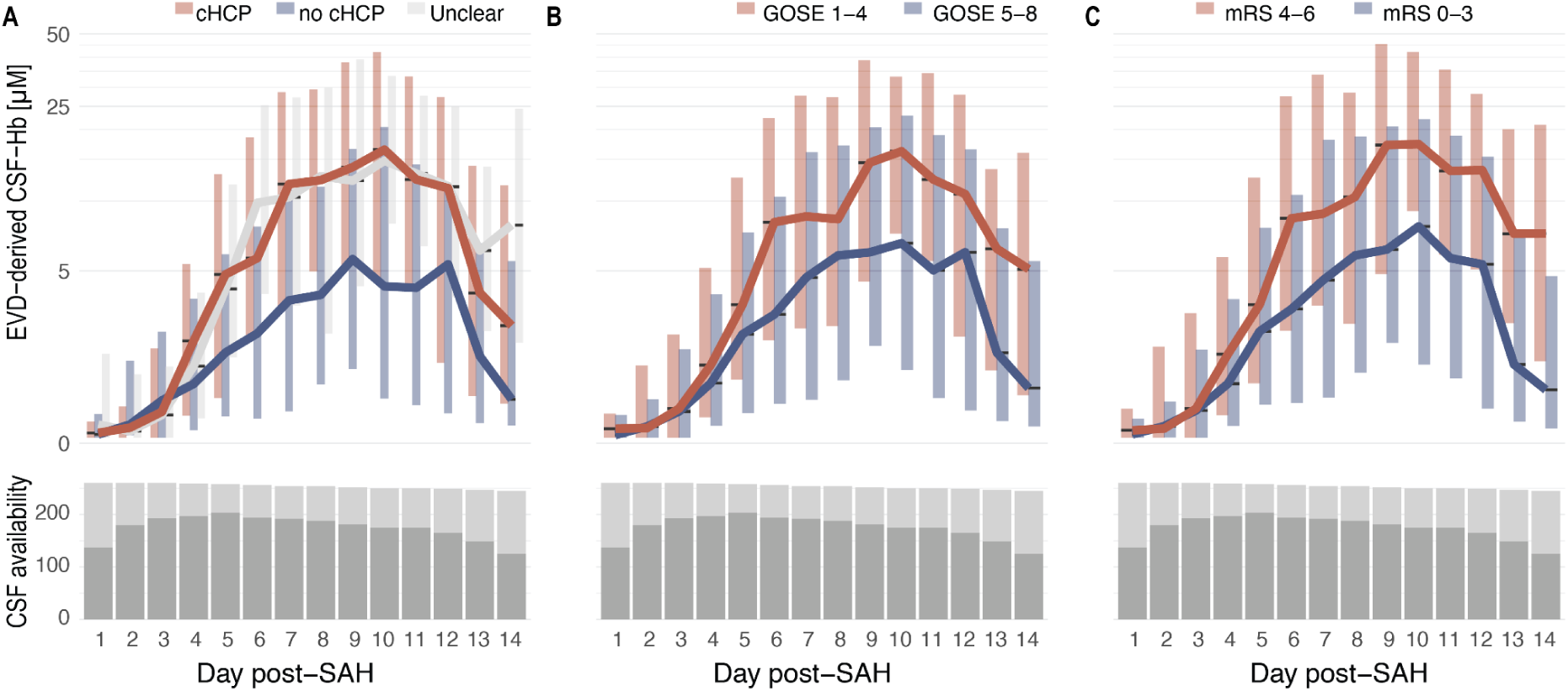
EVD-derived CSF-Hb, chronic hydrocephalus and 3-month outcome. **A.** Acute-phase EVD-derived CSF-Hb stratified by presence versus absence of chronic hydrocephalus. **B.** Acute-phase CSF-Hb levels in patients with favourable (GOSE 5-8) versus unfavourable (GOSE 1-4) 3-month outcome. **C.** Corresponding stratification by modified Rankin Scale (mRS) with favourable (mRS 0-3) versus unfavourable (mRS 4-6) outcome. Bottom panels depict the corresponding availability of CSF samples. *Abbreviations: cHCP, chronic hydrocephalus; CSF-Hb, cerebrospinal fluid oxyhaemoglobin; EVD, external ventricular drain; GOSE, Glasgow Outcome Scale-Extended; mRS, modified Rankin Scale*.

### CSF-metHb and secondary brain injury

As a secondary outcome, we quantified methaemoglobin in CSF (CSF-metHb), the oxidised haemoglobin derivative. EVD-derived CSF-metHb was minimal during the early post-haemorrhagic phase, at or near the lower limit of detection (median 0.1 µM; days 1–5), then rose from day 6 (median 0.123 µM), peaked on day 12 (median 1.41 µM), and declined modestly thereafter while remaining elevated (figure 5A; supplementary table 6).

**Figure 5.**
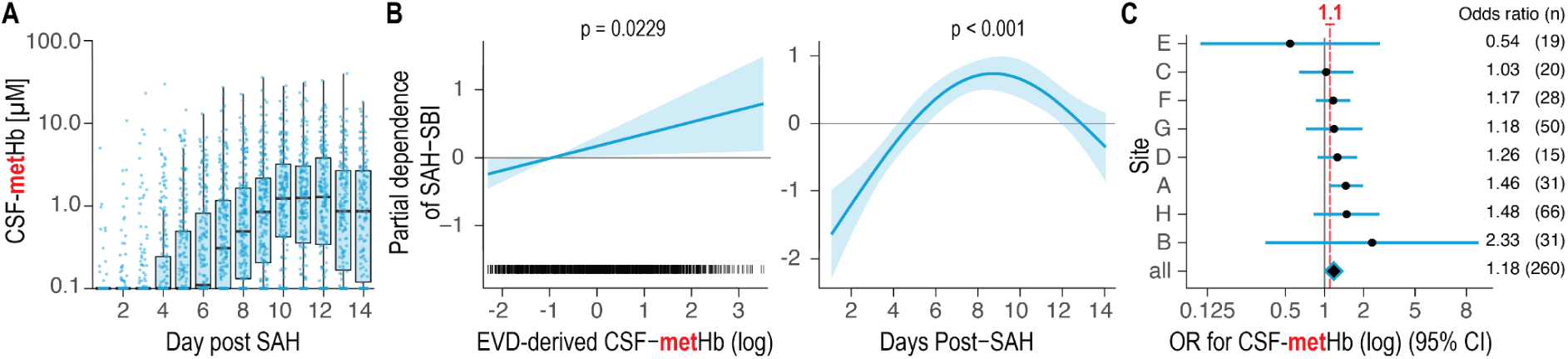
EVD-derived CSF-metHb and secondary brain injury. **A.** Temporal evolution of EVD-derived CSF methaemoglobin (CSF-metHb) during the first 14 days after aneurysmal subarachnoid haemorrhage (aSAH). **B.** Partial-dependence plots from generalised additive models (GAMs) showing a positive association between CSF-metHb and SAH-SBI (p=0.023), with day after haemorrhage as a strong covariate. **C.** Site-specific odds ratios for log-transformed CSF-metHb predicting SAH-SBI, with a pooled odds ratio of 1.18 per log(µM) increase. *Abbreviations: CSF-metHb, cerebrospinal fluid methaemoglobin; SAH-SBI, subarachnoid haemorrhage–related secondary brain injury; OR, odds ratio; GAM, generalised additive model; aSAH, aneurysmal subarachnoid haemorrhage*.

As additional exploratory analysis we analysed CSF-metHb using the same modelling framework as for CSF-Hb. We found that CSF-metHb was significantly associated with SAH-SBI (p=0.023), while day after haemorrhage remained a strong covariate (p<0.001; figure 5B). Site-specific analyses again showed heterogeneity in effect size, but all except one centre demonstrated a positive association between CSF-metHb and SAH-SBI, yielding an overall odds ratio of 1.18 per log(µM) increase in CSF-metHb (figure 5C).

### Safety and procedure-related complications

CSF infection occurred in 2.2% of patients and wound revision in 1.9% (supplementary table 7). No serious study-related events were reported.

## Discussion

### Principal findings

In this preregistered international multicentre study, we evaluated daily EVD-derived CSF-Hb as a real-time monitoring biomarker for SAH-SBI. CSF-Hb showed a characteristic delayed peak on day 10 but had limited prognostic potential for same-day risk stratification: discrimination for SAH-SBI was poor (area under the curve 0.59), and effect estimates varied substantially between centres. In contrast, the oxidised haemoglobin metabolite methaemoglobin was consistently associated with SAH-SBI across sites. Higher acute-phase CSF-Hb concentrations were also related to chronic hydrocephalus and poorer 3-month functional outcome.

Taken together with experimental data,[6–8,15–19] these findings suggest that the limited monitoring performance reflects limitations of a single-analyte, same-day measurement strategy rather than absence of haemolysis-related toxicity. Our results argue against implementing routine CSF-Hb monitoring at the bedside based on single-centre data and underscore the need for rigorous multicentre validation before adopting new neuromonitoring biomarkers.

### Comparison with other studies and implications

The original single-centre study reported a strong association between daily CSF-Hb and SAH-SBI (area under the curve 0.89) and guided our sample size calculation based on the anticipated effect size of OR 0.1 / µM CSF-Hb.[8] In HeMoVal, an effect size in this range was observed at only three centres, predominantly those geographically closest to the pilot site, while other centres showed attenuated or even inverse associations. This pattern likely reflects a combination of regional differences in clinical management, variable sample sizes, and preanalytical variability. Despite harmonised endpoint definitions, between-centre differences in sample sizes, clinical management and assessment of aVSP, DCI, and DIND probably added further noise. The net result was an overall null association between EVD-derived CSF-Hb and SAH-SBI or its individual components.

The observed temporal profiles of CSF-Hb are consistent with earlier single-centre work, but provide more generalisable estimates across diverse practice settings.[6,8] EVD-derived CSF-Hb correlated strongly with baseline haematoma volume and intraventricular haemorrhage, as expected given the larger erythrocyte burden and direct ventricular access. In contrast, these associations were absent or attenuated in lumbar drain samples, probably reflecting haemoglobin degradation, dilution, and compartmental dynamics before blood products reach the lumbar CSF. Temporal patterns also differed by drain type: EVD-derived CSF-Hb showed a delayed rise and peak around day 10, whereas LD-derived CSF-Hb increased earlier and declined after day 10, consistent with compartment-specific erythrocyte breakdown and CSF flow. These findings highlight the importance of sampling location when interpreting CSF biomarkers.

An important practical implication of our findings is safety. Daily manipulation of EVD and LD systems for sampling did not materially increase infection risk: CSF infection rates were low (2.2%) and comparable to reported rates with antimicrobial-coated catheters in routine practice.[20,21] This suggests that frequent sampling can be performed safely under strict aseptic conditions and standardised protocols, which is reassuring for future CSF biomarker studies.

### Strengths and interpretation

Key strengths of this study include its preregistered protocol and statistical analysis plan, multicentre design across eight neurosurgical centres in three countries, consecutive screening of a broad aSAH population, high adherence to daily sampling over 14 days, and blinded central measurement of CSF-Hb and metabolites. The distribution of WFNS grades, aneurysm locations, and treatment modalities supports the clinical representativeness of the cohort.[5,22] The composite SAH-SBI endpoint, integrating aVSP, DCI, and DIND, maximised statistical power, while separate analyses of each component preserved clinical nuance. Generalised additive models allowed flexible modelling of non-linear temporal dynamics and within-patient and within-centre clustering was accounted for using random-effects.

Our analysis was deliberately configured to emulate a potential bedside use case for a single log-transformed analyte (CSF-Hb) used for same-day prediction of SAH-SBI in individual patients. This monitoring-oriented framework is appropriate for testing clinical utility, but conservative for mechanistic inference. It treats cumulative exposure, lagged effects, and coordinated pathway-level patterns as noise rather than signal, and random site effects may absorb biologically meaningful between-centre differences. Mechanistically relevant configurations, such as multi-marker signatures spanning haemolysis, scavenger capacity, inflammation, and neuroglial injury, may therefore be attenuated or entirely missed in this primary analysis.

Nonetheless, several observations support a continued mechanistic role for haemoglobin biology. CSF methaemoglobin was robustly associated with SAH-SBI in almost all centres. First, this discrepancy likely reflects the susceptibility of CSF-Hb to preanalytic erythrocyte lysis, such that measured concentrations may not accurately represent in-vivo CSF-Hb exposure. Second, CSF methaemoglobin represents CSF-Hb that has undergone in-vivo redox conversion and thus captures haemoglobin that has participated in biologically relevant reactions. In addition, higher acute-phase CSF-Hb related to chronic hydrocephalus and unfavourable long-term outcome representing cumulative CSF-Hb burden. These clinical associations are concordant with experimental models in which cell-free haemoglobin and its oxidative products induce vascular dysfunction, oxidative injury, and neuronal damage, and can be mitigated by haemoglobin and heme scavengers.[7,8,16,17,19,23,24] From a therapeutic perspective, our data argue against using CSF-Hb alone as a trigger for intervention, rather than against targeting the haemolytic cascade itself. Effective therapies may need to address upstream haemoglobin toxicity more broadly, encompassing oxyhaemoglobin, oxidised species, and free heme, and be guided by multi-marker, pathway-level signatures rather than a single CSF-Hb concentration.

### Limitations

Several limitations should be considered. First, accurate quantification of CSF-Hb requires meticulous sampling, centrifugation, and handling to avoid pellet contamination and iatrogenic erythrocyte lysis. Despite standard operating procedures and monitoring, residual preanalytical variability between centres is likely and may have diluted associations. CSF-Hb is also labile, undergoing oxidative interconversion in vivo and ex vivo, which can affect measured concentrations. Measurement strategies with minimized preanalytic sample handling, multiple measurements per day, and alternative, more biochemically stable hemolysis biomarkers, such as RBC-specific enzymes, may mitigate these limitations in future studies.

Second, SAH-SBI components (aVSP, DCI, DIND) retain subjective elements and depend on local practice patterns, imaging thresholds, and availability. Even with harmonised definitions and training, inter-observer and inter-institutional variability is unavoidable and probably contributed to heterogeneity in effect estimates. More objective, quantitative endpoints, such as automated vessel calibre assessment, standardised perfusion thresholds, or continuous neuromonitoring metrics, may improve reliability in future multicentre studies.

Third, our primary model intentionally focused on a single marker and same-day outcomes, and did not explicitly model distributed lags, cumulative exposure, or multi-marker patterns. As a result, the negative monitoring result should not be interpreted as exclusion of haemolysis-related mechanisms, which are better addressed in dedicated pathway-level analyses of the HeMoVal biobank.

Finally, the cohort included relatively few participants of non-Caucasian ethnicity, reflecting Central European demographics, which may limit generalisability to more diverse populations.

### Future research

The HeMoVal biobank provides a substantial platform for mechanistic biomarker research and validation of future candidates in aSAH. Beyond single-analyte CSF-Hb, future work will quantify prespecified panels of haemolysis-related proteins, scavenger molecules, inflammatory mediators, and markers of neuroglial injury, and analyse them at the pathway level using longitudinal models that incorporate temporal ordering and distributed lag structures. Such analyses are more closely aligned with the biological hypothesis that haemolysis is an upstream driver of secondary brain injury and may reveal coherent pathway signatures even when individual markers have modest monitoring performance. In parallel, development and clinical testing of automated, near real-time assays for cell-free haemoglobin and its metabolites could reduce preanalytical variability and help clarify whether any haemoglobin-related biomarkers have a role in bedside decision-making.[25]

### Conclusions

In this preregistered international multicentre validation study, EVD-derived CSF-Hb did not perform as a reliable real-time monitoring biomarker for SAH-SBI. Despite a strong mechanistic rationale and promising single-centre data, CSF-Hb showed marked between-centre variability and limited discriminatory ability, arguing against routine clinical implementation of single-analyte CSF-Hb monitoring at the bedside. In contrast, methaemoglobin was associated with secondary brain injury, and higher acute-phase CSF-Hb related to chronic hydrocephalus and poor functional outcome, supporting the relevance of haemoglobin-related toxicity.

Overall, our findings suggest that monitoring CSF oxyhaemoglobin alone is unlikely to be clinically useful to predict SAH-SBI, but highlight the haemolytic cascade as a continuing therapeutic and mechanistic target. Future work using the HeMoVal biobank will apply multi-marker, pathway-level analyses to define haemolysis-related biomarker signatures and provide a platform for robust external validation of future candidates.

## Acknowledgements

We thank the members of the HeMoVal Research Group and we are grateful to the patients for their participation in and commitment to this study.

## Author contributions

Guarantor: DJS.

Concept and design: KA, RMB, DJS, MH.

Obtained funding: KA, RMB, TG, MS, DJS, MH.

Data collection: KA, RMB, EC, AlG, BT, NS, ACH, VK, LB, BEG, JH, GC, NE, AA, CR, LMW, MW, CA, BM, KR, JS, AnG, MG.

Statistical analysis: KA, RMB, BT, DK, UH.

Drafting of the manuscript: KA, RMB.

Critical revision of the manuscript for important intellectual content: all authors.

Administrative, technical, or material support: TG, MS, DC, SW, AW, MI, KB, LR, EK, DJS, MH.

Role of funder: CSL Behring had no role in study design, data collection, interpretation, or the decision to submit for publication.

## HeMoVal Research Group

In addition to the listed authors, the HeMoVal study group further involved:

Zurich, Switzerland: Manuela Grüttner-Durmaz, Susanne Menz, Deborah Chiavi, Rok Humar, Tristan PC van Doormaal.

Aarau, Switzerland: Gerrit A. Schubert, Yasin Irmak. St. Gallen, Switzerland: Oliver Bozinov.

Tuebingen, Germany: Sophie Wang.

Munich, Germany: Ann-Kathrin Jörger, Viktor Maria Eisenkolb

Department of Clinical Research, University of Bern: Muriel Helmers, Danielle Wirz, Dik Heg, Sarina Mahler.

