## Supplemental for "Cerebrospinal fluid haemoglobin as a monitoring biomarker for secondary brain injury after aneurysmal subarachnoid haemorrhage: prospective international multicentre validation study (HeMoVal)"

for the HeMoVal Research Group

*KA and RMB share first authorship; +DJS and MH share last authorship

1 Department of Neurosurgery, Clinical Neuroscience Center, Universitätsspital and University of Zurich, Zurich, Switzerland

2 Division of Internal Medicine, Universitätsspital and University of Zurich, Zurich, Switzerland

3 CSL, CSL Biologics Research Centre, Bern, Switzerland

4 Epidemiology, Biostatistics and Prevention Institute, Department of Biostatistics, University of Zurich, Zurich, Switzerland

5 Department of Neurosurgery, University Hospital Mannheim, University of Heidelberg, Mannheim, Germany

6 Department of Neurosurgery, Aarau Cantonal Hospital, Switzerland

7 Department of Neurosurgery, Technical University of Munich, TUM Medical School, Munich, Germany

8 Department of Neurosurgery, Cantonal Hospital St. Gallen, St. Gallen, Switzerland

9 Department of Neurosurgery, Eberhard Karls University Tuebingen, Tuebingen, Germany

10 Department of Neurosurgery, BG Klinikum Unfallklinik Murnau, Murnau am Staffelsee, Germany

11 Department of Neurosurgery, Kepler University Hospital, Johannes Kepler University Linz, Linz, Austria

12 Clinical Research Institute for Neuroscience, Johannes Kepler University Linz, Linz, Austria

13 Department of Neurosurgery, Comprehensive Center of Clinical Neurosciences and Mental Health, Medical University Vienna, Austria14 Department of Clinical Research, University of Bern, Bern, Switzerland

15 Institute of Neuroradiology, Department of Radiology, Aarau Cantonal Hospital, Switzerland

16 Neurointensive Care Unit, Department of Neurosurgery and Institute of Intensive Care Medicine, Universitätsspital and University of Zurich, Zurich, Switzerland

### Ethical Approvals and Participating Ethics Committees

Ethical approval for this study was obtained from the following institutional review boards and ethics committees:

- Zurich: Kantonale Ethikkommission Zürich (2021-01023)
- Aarau: Ethikkommission Nordwest- und Zentralschweiz (2021-01023)
- St. Gallen: Ethikkommission Ostschweiz (2021-01023)
- Tübingen: Ethikkommission an der Medizinischen Fakultät der Eberhard-Karls-Universität und am Universitätsklinikum Tübingen (758/2021BO1)
- Mannheim: Ethikkommission der Universität Heidelberg (2022-554)
- Munich: Ethikkommission der Technischen Universität München (2022-574-S-KH)
- Vienna: Ethikkommission der Medizinischen Universität Wien (1312/2023)
- Linz: Ethikkommission der Johannes Kepler Universität Linz (1058/2023)

### Supplementary Tables

#### Supplementary Table 1 | Detailed baseline characteristics.

Detailed baseline characteristics of the total (multinational) cohort and the cohorts recruited at the individual study sites categorized in full cohort (F), EVD cohort (EVD) and LD cohort (LD). *Abbreviations:* ACA, anterior cerebral artery; ACOM, Anterior communicating artery; BNI, Barrow Neurological Institute; CAD, coronary artery disease; FND, focal neurological deficit; GCS, Glasgow Coma Scale; MCA, middle cerebral artery; PCOM, posterior communicating artery; PICA, posterior inferior cerebellar artery; WFNS, World Federation of Neurosurgical Societies.

| Characteristic | Site_A_Full | Site_A_EVD | Site_A_LD | Site_B_Full | Site_B_EVD | Site_B_LD | Site_C_Full | Site_C_EVD | Site_C_LD | Site_D_Full | Site_D_EVD | Site_D_LD | Site_E_Full | Site_E_EVD | Site_E_LD | Site_F_Full | Site_F_EVD | Site_F_LD | Site_G_Full | Site_G_EVD | Site_G_LD | Site_H_Full | Site_H_EVD | Site_H_LD |
|---|---|---|---|---|---|---|---|---|---|---|---|---|---|---|---|---|---|---|---|---|---|---|---|---|
| n | 95 | 66 | 20 | 19 | 19 | 0 | 33 | 28 | 3 | 62 | 31 | 10 | 21 | 20 | 8 | 61 | 50 | 1 | 18 | 15 | 3 | 57 | 31 | 21 |
| Age (mean, SD) | 56.7 ± 13.8 | 58.5 ± 13.6 | 59.8 ± 13.1 | 51.6 ± 13.2 | 51.6 ± 13.2 |  | 59.8 ± 11.7 | 60.2 ± 11.8 | 52.3 ± 12.6 | 58.2 ± 14.4 | 63 ± 13.9 | 58.8 ± 16.3 | 60.8 ± 12.8 | 61.4 ± 12.9 | 55 ± 9.4 | 57.7 ± 11.8 | 57.9 ± 12.5 | 58 ± NA | 60.8 ± 10.3 | 62.1 ± 10.8 | 54.3 ± 2.9 | 59.5 ± 13.6 | 61.8 ± 15.1 | 56.8 ± 9.9 |
| Male sex (n, %) | 34 (35.8%) | 19 (28.8%) | 6 (30%) | 7 (36.8%) | 7 (36.8%) |  | 9 (27.3%) | 8 (28.6%) | 1 (33.3%) | 22 (35.5%) | 8 (25.8%) | 4 (40%) | 6 (28.6%) | 5 (25%) | 3 (37.5%) | 28 (45.9%) | 23 (46%) | 1 (100%) | 5 (27.8%) | 4 (26.7%) | 1 (33.3%) | 17 (29.8%) | 8 (25.8%) | 7 (33.3%) |
| **Diabetes mellitus (n, %)** | 2 (2.1%) | 2 (3%) | 0 (0%) | 2 (10.5%) | 2 (10.5%) |  | 2 (6.1%) | 2 (7.1%) | 0 (0%) | 0 (0%) | 0 (0%) | 0 (0%) | 1 (4.8%) | 1 (5%) | 0 (0%) | 2 (3.3%) | 2 (4%) | 0 (0%) | 0 (0%) | 0 (0%) | 0 (0%) | 3 (5.3%) | 0 (0%) | 3 (14.3%) |
| Hypertension (n, %) | 29 (30.5%) | 21 (31.8%) | 8 (40%) | 9 (47.4%) | 9 (47.4%) |  | 14 (42.4%) | 11 (39.3%) | 1 (33.3%) | 15 (24.2%) | 7 (22.6%) | 1 (10%) | 11 (52.4%) | 10 (50%) | 3 (37.5%) | 33 (54.1%) | 25 (50%) | 1 (100%) | 7 (38.9%) | 7 (46.7%) | 0 (0%) | 17 (29.8%) | 8 (25.8%) | 7 (33.3%) |
| CAD (n, %) | 5 (5.3%) | 3 (4.5%) | 2 (10%) | 1 (5.3%) | 1 (5.3%) |  | 1 (3%) | 1 (3.6%) | 0 (0%) | 5 (8.1%) | 2 (6.5%) | 0 (0%) | 3 (14.3%) | 3 (15%) | 0 (0%) | 12 (19.7%) | 11 (22%) | 0 (0%) | 2 (11.1%) | 2 (13.3%) | 0 (0%) | 5 (8.8%) | 3 (9.7%) | 2 (9.5%) |
| **Antithrombotics (n, %)** |  |  |  |  |  |  |  |  |  |  |  |  |  |  |  |  |  |  |  |  |  |  |  |  |
| None | 82 (86.3%) | 58 (87.9%) | 15 (75%) | 15 (78.9%) | 15 (78.9%) |  | 29 (87.9%) | 24 (85.7%) | 3 (100%) | 50 (80.6%) | 22 (71%) | 7 (70%) | 16 (76.2%) | 15 (75%) | 8 (100%) | 49 (80.3%) | 39 (78%) | 1 (100%) | 16 (88.9%) | 13 (86.7%) | 3 (100%) | 46 (80.7%) | 25 (80.6%) | 17 (81%) |
| AP | 8 (8.4%) | 5 (7.6%) | 3 (15%) | 1 (5.3%) | 1 (5.3%) |  | 2 (6.1%) | 2 (7.1%) |  | 6 (9.7%) | 4 (12.9%) | 2 (20%) |  |  |  | 10 (16.4%) | 9 (18%) |  |  |  |  | 6 (10.5%) | 4 (12.9%) | 2 (9.5%) |
| AC | 5 (5.3%) | 3 (4.5%) | 2 (10%) | 2 (10.5%) | 2 (10.5%) |  | 1 (3%) | 1 (3.6%) |  | 1 (1.6%) | 1 (3.2%) |  | 1 (4.8%) | 1 (5%) |  | 1 (1.6%) | 1 (2%) |  | 1 (5.6%) | 1 (6.7%) |  | 5 (8.8%) | 2 (6.5%) | 2 (9.5%) |
| **Smoking (n, %)** |  |  |  |  |  |  |  |  |  |  |  |  |  |  |  |  |  |  |  |  |  |  |  |  |
| Never | 36 (37.9%) | 24 (36.4%) | 6 (30%) | 2 (10.5%) | 2 (10.5%) |  | 2 (6.1%) | 2 (7.1%) |  | 21 (33.9%) | 8 (25.8%) | 3 (30%) | 3 (14.3%) | 3 (15%) |  | 14 (23%) | 13 (26%) |  | 4 (22.2%) | 2 (13.3%) | 2 (66.7%) | 7 (12.3%) | 4 (12.9%) | 1 (4.8%) |
| Patient is a current smoker. | 27 (28.4%) | 15 (22.7%) | 5 (25%) | 7 (36.8%) | 7 (36.8%) |  | 7 (21.2%) | 6 (21.4%) | 1 (33.3%) | 22 (35.5%) | 12 (38.7%) | 2 (20%) | 3 (14.3%) | 2 (10%) | 3 (37.5%) | 21 (34.4%) | 15 (30%) | 1 (100%) | 5 (27.8%) | 4 (26.7%) | 1 (33.3%) | 21 (36.8%) | 8 (25.8%) | 9 (42.9%) |
| Patient quit smoking. | 1 (1.1%) | 1 (1.5%) | 1 (5%) | 1 (5.3%) | 1 (5.3%) |  | 2 (6.1%) | 2 (7.1%) |  | 3 (4.8%) | 2 (6.5%) |  |  |  |  | 4 (6.6%) | 2 (4%) |  | 4 (22.2%) | 4 (26.7%) |  | 1 (1.8%) | 1 (3.2%) |  |
| Missing data | 31 (32.6%) | 26 (39.4%) | 8 (40%) | 9 (47.4%) | 9 (47.4%) |  | 22 (66.7%) | 18 (64.3%) | 2 (66.7%) | 16 (25.8%) | 9 (29%) | 5 (50%) | 15 (71.4%) | 15 (75%) | 5 (62.5%) | 22 (36.1%) | 20 (40%) |  | 5 (27.8%) | 5 (33.3%) |  | 28 (49.1%) | 18 (58.1%) | 11 (52.4%) |
| **GCS (mean, SD)** | 12.9 ± 2.6 | 11.7 ± 2.9 | 12.8 ± 1.9 | 14.5 ± 0.7 | 14.5 ± 0.7 |  | 13.6 ± 3.2 | 13.1 ± 3.5 | 15 ± 0 | 13.9 ± 1.7 | 13.3 ± 1.7 | 13.7 ± 1.9 | 11.4 ± 4.8 | 11 ± 4.9 | 13.4 ± 3 | 13.6 ± 2.5 | 13.2 ± 2.8 | NA | 14.1 ± 0.9 | 14.1 ± 0.9 | 14 ± 1 | 13.9 ± 2 | 12.6 ± 2.6 | 14.1 ± 2 |
| **Pupil status (n, %)** |  |  |  |  |  |  |  |  |  |  |  |  |  |  |  |  |  |  |  |  |  |  |  |  |
| no pathological mydriasis. | 82 (86.3%) | 58 (87.9%) | 18 (90%) | 18 (94.7%) | 18 (94.7%) |  | 23 (69.7%) | 19 (67.9%) | 2 (66.7%) | 57 (91.9%) | 30 (96.8%) | 8 (80%) | 16 (76.2%) | 15 (75%) | 6 (75%) | 49 (80.3%) | 39 (78%) |  | 18 (100%) | 15 (100%) | 3 (100%) | 50 (87.7%) | 25 (80.6%) | 21 (100%) |
| Anisocoria | 12 (12.6%) | 8 (12.1%) | 1 (5%) |  |  |  | 3 (9.1%) | 2 (7.1%) |  | 4 (6.5%) | 1 (3.2%) | 1 (10%) | 1 (4.8%) | 1 (5%) |  | 3 (4.9%) | 3 (6%) |  |  |  |  | 7 (12.3%) | 6 (19.4%) |  |
| Bilateral mydriasis |  |  |  |  |  |  |  |  |  | 1 (1.6%) |  | 1 (10%) |  |  |  | 1 (1.6%) | 1 (2%) |  |  |  |  |  |  |  |
| Missing data | 1 (1.1%) |  | 1 (5%) | 1 (5.3%) | 1 (5.3%) |  | 7 (21.2%) | 7 (25%) | 1 (33.3%) |  |  |  | 4 (19%) | 4 (20%) | 2 (25%) | 8 (13.1%) | 7 (14%) | 1 (100%) |  |  |  |  |  |  |
| **Cranial nerve deficit (n, %)** | 5 (5.3%) | 3 (4.5%) | 2 (10%) | 0 (0%) | 0 (0%) |  | 2 (6.1%) | 2 (7.1%) | 0 (0%) | 5 (8.1%) | 1 (3.2%) | 0 (0%) | 0 (0%) | 0 (0%) | 0 (0%) | 3 (4.9%) | 3 (6%) | 0 (0%) | 0 (0%) | 0 (0%) | 0 (0%) | 1 (1.8%) | 1 (3.2%) | 0 (0%) |
| **Focal neurological deficit (n, %)** |  |  |  |  |  |  |  |  |  |  |  |  |  |  |  |  |  |  |  |  |  |  |  |  |
| No | 47 (49.5%) | 19 (28.8%) | 10 (50%) | 11 (57.9%) | 11 (57.9%) |  | 18 (54.5%) | 14 (50%) | 2 (66.7%) | 45 (72.6%) | 18 (58.1%) | 8 (80%) | 8 (38.1%) | 7 (35%) | 4 (50%) | 32 (52.5%) | 22 (44%) |  | 9 (50%) | 8 (53.3%) | 1 (33.3%) | 35 (61.4%) | 12 (38.7%) | 17 (81%) |
| Motor | 5 (5.3%) | 5 (7.6%) | 2 (10%) | 1 (5.3%) | 1 (5.3%) |  |  |  |  | 3 (4.8%) | 2 (6.5%) |  | 1 (4.8%) | 1 (5%) |  | 5 (8.2%) | 5 (10%) |  | 1 (5.6%) |  | 1 (33.3%) | 1 (1.8%) | 1 (3.2%) |  |
| Sensory |  |  |  |  |  |  |  |  |  |  |  |  |  |  |  |  |  |  |  |  |  | 1 (1.8%) |  |  |
| Motor and sensory | 2 (2.1%) | 2 (3%) | 1 (5%) | 1 (5.3%) | 1 (5.3%) |  |  |  |  | 1 (1.6%) | 1 (3.2%) |  |  |  |  | 1 (1.6%) | 1 (2%) |  |  |  |  |  |  |  |
| Missing data | 7 (7.4%) | 7 (10.6%) |  |  |  |  | 1 (3%) | 1 (3.6%) |  | 1 (1.6%) |  | 1 (10%) | 2 (9.5%) | 2 (10%) | 1 (12.5%) | 1 (1.6%) | 1 (2%) |  | 3 (16.7%) | 2 (13.3%) | 1 (33.3%) | 1 (1.8%) | 1 (3.2%) |  |
| **Headache (n, %)** |  |  |  |  |  |  |  |  |  |  |  |  |  |  |  |  |  |  |  |  |  |  |  |  |
| No | 5 (5.3%) | 2 (3%) | 2 (10%) |  |  |  | 2 (6.1%) | 2 (7.1%) |  | 6 (9.7%) | 1 (3.2%) |  |  |  |  | 3 (4.9%) | 3 (6%) |  |  |  |  | 2 (3.5%) |  |  |
| Minimal | 3 (3.2%) | 1 (1.5%) |  | 1 (5.3%) | 1 (5.3%) |  |  |  |  | 3 (4.8%) | 1 (3.2%) |  |  |  |  | 3 (4.9%) | 2 (4%) |  | 1 (5.6%) | 1 (6.7%) |  | 5 (8.8%) |  | 4 (19%) |
| Moderate | 7 (7.4%) | 4 (6.1%) | 1 (5%) | 3 (15.8%) | 3 (15.8%) |  | 8 (24.2%) | 6 (21.4%) | 2 (66.7%) | 8 (12.9%) | 3 (9.7%) | 1 (10%) | 2 (9.5%) | 2 (10%) |  | 11 (18%) | 9 (18%) |  | 3 (16.7%) | 2 (13.3%) | 1 (33.3%) | 11 (19.3%) | 5 (16.1%) | 3 (14.3%) |
| Severe | 37 (38.9%) | 18 (27.3%) | 8 (40%) | 9 (47.4%) | 9 (47.4%) |  | 7 (21.2%) | 5 (17.9%) |  | 29 (46.8%) | 13 (41.9%) | 8 (80%) | 7 (33.3%) | 6 (30%) | 4 (50%) | 21 (34.4%) | 14 (28%) |  | 8 (44.4%) | 7 (46.7%) | 1 (33.3%) | 18 (31.6%) | 8 (25.8%) | 9 (42.9%) |
| Missing data | 9 (9.5%) | 8 (12.1%) | 2 (10%) |  |  |  | 2 (6.1%) | 2 (7.1%) |  | 4 (6.5%) | 3 (9.7%) |  | 2 (9.5%) | 2 (10%) | 1 (12.5%) | 1 (1.6%) | 1 (2%) |  | 1 (5.6%) |  | 1 (33.3%) | 2 (3.5%) | 1 (3.2%) | 1 (4.8%) |
| **Neck stiffness (n, %)** |  |  |  |  |  |  |  |  |  |  |  |  |  |  |  |  |  |  |  |  |  |  |  |  |
| No | 25 (26.3%) | 13 (19.7%) | 7 (35%) | 10 (52.6%) | 10 (52.6%) |  | 9 (27.3%) | 6 (21.4%) | 1 (33.3%) | 25 (40.3%) | 10 (32.3%) | 4 (40%) | 3 (14.3%) | 3 (15%) | 1 (12.5%) | 7 (11.5%) | 4 (8%) |  | 3 (16.7%) | 3 (20%) |  | 18 (31.6%) | 3 (9.7%) | 10 (47.6%) |
| Slight | 9 (9.5%) | 3 (4.5%) | 1 (5%) |  |  |  | 4 (12.1%) | 3 (10.7%) |  | 17 (27.4%) | 7 (22.6%) | 3 (30%) | 3 (14.3%) | 3 (15%) | 2 (25%) | 11 (18%) | 8 (16%) |  | 3 (16.7%) | 2 (13.3%) | 1 (33.3%) | 13 (22.8%) | 8 (25.8%) | 4 (19%) |
| Pronounced | 14 (14.7%) | 5 (7.6%) | 2 (10%) | 1 (5.3%) | 1 (5.3%) |  | 3 (9.1%) | 3 (10.7%) | 1 (33.3%) | 3 (4.8%) | 2 (6.5%) |  | 1 (4.8%) | 1 (5%) |  | 13 (21.3%) | 9 (18%) |  | 2 (11.1%) | 1 (6.7%) | 1 (33.3%) | 1 (1.8%) | 1 (3.2%) |  |
| Missing data | 13 (13.7%) | 12 (18.2%) | 3 (15%) | 2 (10.5%) | 2 (10.5%) |  | 3 (9.1%) | 3 (10.7%) |  | 5 (8.1%) | 2 (6.5%) | 2 (20%) | 4 (19%) | 3 (15%) | 2 (25%) | 8 (13.1%) | 8 (16%) |  | 5 (27.8%) | 4 (26.7%) | 1 (33.3%) | 6 (10.5%) | 2 (6.5%) | 3 (14.3%) |
| **WFNS grade (n, %)** |  |  |  |  |  |  |  |  |  |  |  |  |  |  |  |  |  |  |  |  |  |  |  |  |
| 1 | 19 (20%) | 3 (4.5%) | 2 (10%) | 8 (42.1%) | 8 (42.1%) |  | 14 (42.4%) | 9 (32.1%) | 2 (66.7%) | 27 (43.5%) | 5 (16.1%) | 3 (30%) | 8 (38.1%) | 7 (35%) | 4 (50%) | 20 (32.8%) | 12 (24%) |  | 5 (27.8%) | 4 (26.7%) | 1 (33.3%) | 23 (40.4%) | 3 (9.7%) | 11 (52.4%) |
| 2 | 26 (27.4%) | 16 (24.2%) | 8 (40%) | 4 (21.1%) | 4 (21.1%) |  | 3 (9.1%) | 3 (10.7%) |  | 16 (25.8%) | 12 (38.7%) | 6 (60%) | 4 (19%) | 4 (20%) | 1 (12.5%) | 12 (19.7%) | 10 (20%) |  | 7 (38.9%) | 7 (46.7%) |  | 13 (22.8%) | 10 (32.3%) | 5 (23.8%) |
| 3 | 5 (5.3%) | 4 (6.1%) | 2 (10%) | 2 (10.5%) | 2 (10.5%) |  |  |  |  | 1 (1.6%) | 1 (3.2%) |  |  |  |  | 2 (3.3%) | 2 (4%) |  | 3 (16.7%) | 1 (6.7%) | 2 (66.7%) |  |  |  |
| 4 | 23 (24.2%) | 21 (31.8%) | 5 (25%) | 1 (5.3%) | 1 (5.3%) |  | 6 (18.2%) | 6 (21.4%) | 1 (33.3%) | 12 (19.4%) | 8 (25.8%) | 1 (10%) | 3 (14.3%) | 3 (15%) | 1 (12.5%) | 15 (24.6%) | 15 (30%) |  | 1 (5.6%) | 1 (6.7%) |  | 13 (22.8%) | 10 (32.3%) | 4 (19%) |
| 5 | 22 (23.2%) | 22 (33.3%) | 3 (15%) | 4 (21.1%) | 4 (21.1%) |  | 10 (30.3%) | 10 (35.7%) |  | 6 (9.7%) | 5 (16.1%) |  | 6 (28.6%) | 6 (30%) | 2 (25%) | 12 (19.7%) | 11 (22%) | 1 (100%) | 2 (11.1%) | 2 (13.3%) |  | 8 (14%) | 8 (25.8%) | 1 (4.8%) |
| **Hunt & Hess grade (n, %)** |  |  |  |  |  |  |  |  |  |  |  |  |  |  |  |  |  |  |  |  |  |  |  |  |
| 1 | 2 (2.1%) | 1 (1.5%) | 1 (5%) | 2 (10.5%) | 2 (10.5%) |  | 1 (3%) |  |  | 2 (3.2%) |  |  |  |  |  | 4 (6.6%) | 3 (6%) |  |  |  |  | 5 (8.8%) |  | 3 (14.3%) |
| 2 | 35 (36.8%) | 10 (15.2%) | 8 (40%) | 9 (47.4%) | 9 (47.4%) |  | 15 (45.5%) | 12 (42.9%) | 2 (66.7%) | 39 (62.9%) | 16 (51.6%) | 9 (90%) | 8 (38.1%) | 7 (35%) | 4 (50%) | 28 (45.9%) | 19 (38%) |  | 9 (50%) | 8 (53.3%) | 1 (33.3%) | 25 (43.9%) | 9 (29%) | 9 (42.9%) |
| 3 | 25 (26.3%) | 22 (33.3%) | 4 (20%) | 5 (26.3%) | 5 (26.3%) |  | 6 (18.2%) | 5 (17.9%) | 1 (33.3%) | 9 (14.5%) | 7 (22.6%) |  | 3 (14.3%) | 3 (15%) | 1 (12.5%) | 10 (16.4%) | 10 (20%) |  | 6 (33.3%) | 4 (26.7%) | 2 (66.7%) | 11 (19.3%) | 7 (22.6%) | 6 (28.6%) |
| 4 | 16 (16.8%) | 16 (24.2%) | 4 (20%) | 1 (5.3%) | 1 (5.3%) |  | 6 (18.2%) | 6 (21.4%) |  | 8 (12.9%) | 4 (12.9%) | 1 (10%) | 8 (38.1%) | 8 (40%) | 3 (37.5%) | 15 (24.6%) | 15 (30%) |  | 3 (16.7%) | 3 (20%) |  | 13 (22.8%) | 12 (38.7%) | 3 (14.3%) |
| 5 | 17 (17.9%) | 17 (25.8%) | 3 (15%) | 2 (10.5%) | 2 (10.5%) |  | 5 (15.2%) | 5 (17.9%) |  | 4 (6.5%) | 4 (12.9%) |  | 2 (9.5%) | 2 (10%) |  | 4 (6.6%) | 3 (6%) | 1 (100%) |  |  |  | 3 (5.3%) | 3 (9.7%) |  |
| **Aneurysm location (n, %)** |  |  |  |  |  |  |  |  |  |  |  |  |  |  |  |  |  |  |  |  |  |  |  |  |
| ACA | 6 (6.3%) | 5 (7.6%) | 1 (5%) | 1 (5.3%) | 1 (5.3%) |  | 1 (3%) | 1 (3.6%) |  | 4 (6.5%) | 2 (6.5%) | 1 (10%) | 1 (4.8%) | 1 (5%) | 1 (12.5%) | 2 (3.3%) | 1 (2%) |  | 1 (5.6%) |  | 1 (33.3%) | 5 (8.8%) | 3 (9.7%) | 1 (4.8%) |
| ACOM | 31 (32.6%) | 20 (30.3%) | 10 (50%) | 10 (52.6%) | 10 (52.6%) |  | 12 (36.4%) | 11 (39.3%) | 1 (33.3%) | 27 (43.5%) | 12 (38.7%) | 6 (60%) | 10 (47.6%) | 10 (50%) | 4 (50%) | 18 (29.5%) | 14 (28%) |  | 8 (44.4%) | 7 (46.7%) | 1 (33.3%) | 14 (24.6%) | 8 (25.8%) | 5 (23.8%) |
| AntCh | 1 (1.1%) | 1 (1.5%) |  |  |  |  |  |  |  |  |  |  |  |  |  | 3 (4.9%) | 2 (4%) |  |  |  |  |  |  |  |
| B_o | 1 (1.1%) | 1 (1.5%) |  |  |  |  |  |  |  |  |  |  |  |  |  | 1 (1.6%) | 1 (2%) |  | 1 (5.6%) | 1 (6.7%) |  | 1 (1.8%) | 1 (3.2%) |  |
| B_t | 11 (11.6%) | 10 (15.2%) | 4 (20%) | 1 (5.3%) | 1 (5.3%) |  | 1 (3%) | 1 (3.6%) |  | 3 (4.8%) | 3 (9.7%) | 1 (10%) |  |  |  | 3 (4.9%) | 2 (4%) | 1 (100%) | 2 (11.1%) | 1 (6.7%) | 1 (33.3%) | 3 (5.3%) | 1 (3.2%) | 1 (4.8%) |
| ICA_o | 9 (9.5%) | 6 (9.1%) | 1 (5%) |  |  |  | 3 (9.1%) | 2 (7.1%) | 1 (33.3%) | 1 (1.6%) | 1 (3.2%) |  |  |  |  | 4 (6.6%) | 3 (6%) |  |  |  |  | 3 (5.3%) | 2 (6.5%) |  |
| ICA_po | 2 (2.1%) | 2 (3%) |  |  |  |  |  |  |  | 5 (8.1%) | 2 (6.5%) | 1 (10%) |  |  |  | 2 (3.3%) | 1 (2%) |  | 1 (5.6%) | 1 (6.7%) |  |  |  |  |
| MCA | 15 (15.8%) | 7 (10.6%) | 2 (10%) | 3 (15.8%) | 3 (15.8%) |  | 6 (18.2%) | 3 (10.7%) |  | 13 (21%) | 4 (12.9%) | 1 (10%) | 8 (38.1%) | 7 (35%) | 3 (37.5%) | 14 (23%) | 13 (26%) |  | 3 (16.7%) | 3 (20%) |  | 12 (21.1%) | 3 (9.7%) | 8 (38.1%) |
| PCA | 2 (2.1%) | 1 (1.5%) |  |  |  |  |  |  |  |  |  |  |  |  |  |  |  |  |  |  |  | 3 (5.3%) | 2 (6.5%) |  |
| PCOM | 11 (11.6%) | 7 (10.6%) |  | 2 (10.5%) | 2 (10.5%) |  | 4 (12.1%) | 4 (14.3%) | 1 (33.3%) | 6 (9.7%) | 4 (12.9%) |  | 2 (9.5%) | 2 (10%) |  | 6 (9.8%) | 5 (10%) |  |  |  |  | 10 (17.5%) | 6 (19.4%) | 4 (19%) |
| PICA | 3 (3.2%) | 3 (4.5%) | 1 (5%) | 1 (5.3%) | 1 (5.3%) |  | 2 (6.1%) | 2 (7.1%) |  | 2 (3.2%) | 2 (6.5%) |  |  |  |  | 6 (9.8%) | 6 (12%) |  | 1 (5.6%) | 1 (6.7%) |  | 3 (5.3%) | 3 (9.7%) | 1 (4.8%) |
| SCA | 1 (1.1%) | 1 (1.5%) | 1 (5%) |  |  |  | 1 (3%) | 1 (3.6%) |  |  |  |  |  |  |  |  |  |  |  |  |  |  |  |  |
| VA | 2 (2.1%) | 2 (3%) |  | 1 (5.3%) | 1 (5.3%) |  | 3 (9.1%) | 3 (10.7%) |  | 1 (1.6%) | 1 (3.2%) |  |  |  |  | 2 (3.3%) | 2 (4%) |  | 1 (5.6%) | 1 (6.7%) |  | 3 (5.3%) | 2 (6.5%) | 1 (4.8%) |
| **Aneurysm size (mean, SD)** | 8.7 ± 9.2 | 8.9 ± 9.9 | 6.2 ± 3.3 | 6 ± 4.7 | 6 ± 4.7 |  | 6.7 ± 4.2 | 7.1 ± 4.3 | 5 ± 1 | 7.4 ± 9.2 | 6.5 ± 3.7 | 6.5 ± 3.3 | 7.2 ± 3.5 | 7.2 ± 3.5 | 5.9 ± 0.4 | 4.9 ± 3.4 | 4.7 ± 3.3 | 10 ± NA | 4.7 ± 2.5 | 5 ± 2.6 | 3.3 ± 1.5 | 7.5 ± 4.2 | 7.6 ± 4.3 | 6.7 ± 3.7 |
| **Intracerebral hemorrhage (n, %)** | 20 (21.1%) | 17 (25.8%) | 4 (20%) | 8 (42.1%) | 8 (42.1%) |  | 22 (66.7%) | 18 (64.3%) | 2 (66.7%) | 13 (21%) | 6 (19.4%) | 0 (0%) | 15 (71.4%) | 15 (75%) | 6 (75%) | 24 (39.3%) | 22 (44%) | 0 (0%) | 4 (22.2%) | 4 (26.7%) | 0 (0%) | 21 (36.8%) | 13 (41.9%) | 9 (42.9%) |
| **Intraventricular hemorrhage (n, %)** | 54 (56.8%) | 47 (71.2%) | 11 (55%) | 9 (47.4%) | 9 (47.4%) |  | 18 (54.5%) | 18 (64.3%) | 1 (33.3%) | 35 (56.5%) | 26 (83.9%) | 8 (80%) | 16 (76.2%) | 16 (80%) | 6 (75%) | 39 (63.9%) | 35 (70%) | 1 (100%) | 9 (50%) | 8 (53.3%) | 1 (33.3%) | 17 (29.8%) | 15 (48.4%) | 2 (9.5%) |
| **Modified Fisher grade (n, %)** |  |  |  |  |  |  |  |  |  |  |  |  |  |  |  |  |  |  |  |  |  |  |  |  |
| 1 | 11 (11.6%) | 4 (6.1%) | 2 (10%) | 4 (21.1%) | 4 (21.1%) |  | 3 (9.1%) |  |  | 14 (22.6%) | 1 (3.2%) | 2 (20%) | 1 (4.8%) |  | 1 (12.5%) | 7 (11.5%) | 3 (6%) |  | 4 (22.2%) | 3 (20%) | 1 (33.3%) | 15 (26.3%) | 3 (9.7%) | 5 (23.8%) |
| 2 | 3 (3.2%) | 2 (3%) |  | 2 (10.5%) | 2 (10.5%) |  | 4 (12.1%) | 4 (14.3%) |  | 7 (11.3%) | 4 (12.9%) | 3 (30%) | 1 (4.8%) | 1 (5%) |  | 5 (8.2%) | 5 (10%) |  | 3 (16.7%) | 3 (20%) |  | 4 (7%) | 2 (6.5%) | 2 (9.5%) |
| 3 | 30 (31.6%) | 15 (22.7%) | 7 (35%) | 6 (31.6%) | 6 (31.6%) |  | 12 (36.4%) | 10 (35.7%) | 2 (66.7%) | 13 (21%) | 4 (12.9%) |  | 4 (19%) | 4 (20%) | 1 (12.5%) | 15 (24.6%) | 12 (24%) |  | 5 (27.8%) | 4 (26.7%) | 1 (33.3%) | 25 (43.9%) | 13 (41.9%) | 14 (66.7%) |
| 4 | 51 (53.7%) | 45 (68.2%) | 11 (55%) | 7 (36.8%) | 7 (36.8%) |  | 14 (42.4%) | 14 (50%) | 1 (33.3%) | 28 (45.2%) | 22 (71%) | 5 (50%) | 15 (71.4%) | 15 (75%) | 6 (75%) | 34 (55.7%) | 30 (60%) | 1 (100%) | 6 (33.3%) | 5 (33.3%) | 1 (33.3%) | 13 (22.8%) | 13 (41.9%) |  |
| **BNI grade (n, %)** |  |  |  |  |  |  |  |  |  |  |  |  |  |  |  |  |  |  |  |  |  |  |  |  |
| No visible SAH |  |  |  | 1 (5.3%) | 1 (5.3%) |  | 2 (6.1%) | 1 (3.6%) |  | 3 (4.8%) |  | 1 (10%) |  |  |  | 1 (1.6%) |  |  |  |  |  | 2 (3.5%) |  |  |
| Maximum SAH thickness | 20 (21.1%) | 19 (28.8%) | 4 (20%) | 3 (15.8%) | 3 (15.8%) |  | 7 (21.2%) | 7 (25%) | 2 (66.7%) | 2 (3.2%) | 2 (6.5%) | 2 (20%) | 6 (28.6%) | 6 (30%) | 2 (25%) | 10 (16.4%) | 9 (18%) | 1 (100%) | 2 (11.1%) | 1 (6.7%) | 1 (33.3%) | 9 (15.8%) | 8 (25.8%) | 2 (9.5%) |
| **Hematoma volume (mean, SD)** | 61.3 ± 39.3 | 61.3 ± 39.3 | 56.9 ± 36.9 | 69.9 ± 69.2 | 69.9 ± 69.2 |  | 38.5 ± 26.5 | 38.5 ± 26.5 | 28.4 ± NA | 40.9 ± 27.8 | 40.9 ± 27.8 | 33.5 ± 30.8 | 80.1 ± 56.2 | 80.1 ± 56.2 | 91 ± 73.5 | 54.5 ± NA | 54.5 ± NA | NA | NA | NA | NA | 80.7 ± 96.3 | 80.7 ± 96.3 | 166.3 ± 222.2 |
| **Aneurysm therapy (n, %)** |  |  |  |  |  |  |  |  |  |  |  |  |  |  |  |  |  |  |  |  |  |  |  |  |
| Bypass |  |  |  |  |  |  |  |  |  |  |  |  |  |  |  | 2 (3.3%) | 2 (4%) |  |  |  |  | 1 (1.8%) |  |  |
| Clipping | 33 (34.7%) | 16 (24.2%) | 6 (30%) | 2 (10.5%) | 2 (10.5%) |  | 16 (48.5%) | 12 (42.9%) |  | 13 (21%) | 3 (9.7%) | 1 (10%) | 11 (52.4%) | 11 (55%) | 4 (50%) | 32 (52.5%) | 28 (56%) |  | 5 (27.8%) | 5 (33.3%) |  | 22 (38.6%) | 8 (25.8%) | 11 (52.4%) |
| Coiling | 58 (61.1%) | 46 (69.7%) | 13 (65%) | 16 (84.2%) | 16 (84.2%) |  | 16 (48.5%) | 15 (53.6%) | 3 (100%) | 45 (72.6%) | 25 (80.6%) | 8 (80%) | 4 (19%) | 4 (20%) |  | 23 (37.7%) | 16 (32%) | 1 (100%) | 10 (55.6%) | 9 (60%) | 1 (33.3%) | 32 (56.1%) | 21 (67.7%) | 10 (47.6%) |
| Flow diverter | 4 (4.2%) | 4 (6.1%) | 1 (5%) |  |  |  |  |  |  | 2 (3.2%) | 2 (6.5%) |  | 3 (14.3%) | 2 (10%) | 1 (12.5%) | 4 (6.6%) | 4 (8%) |  | 3 (16.7%) | 1 (6.7%) | 2 (66.7%) |  |  |  |
| Others |  |  |  | 1 (5.3%) | 1 (5.3%) |  | 1 (3%) | 1 (3.6%) |  | 2 (3.2%) | 1 (3.2%) | 1 (10%) | 3 (14.3%) | 3 (15%) | 3 (37.5%) |  |  |  |  |  |  |  |  |  |
| None |  |  |  |  |  |  |  |  |  |  |  |  |  |  |  |  |  |  |  |  |  | 2 (3.5%) | 2 (6.5%) |  |

**Supplementary Table 2 | Daily CSF-Hb levels within 14 days of aneurysm rupture.**

The median, mean and the SD of CSF-Hb measurements (in μM) within the total (multicenter) EVD and LD cohorts stratified by day post-SAH. The difference in mean EVD-derived and LD-derived CSF-Hb is given with the corresponding 95% confidence interval and p-value (Mann-Whitney U test).

| **Day post-SAH** | **EVD Cohort** | | **LD cohort** | | **Difference in mean with 95% CI** |
|---|---|---|---|---|---|
| | *median* | *mean (SD)* | *median* | *mean (SD)* | *difference [95%CI] (p value)* |
| 1 | 0.188 | 12.547 (94.461) | 0.325 | 46.827 (219.219) | -34.28 [-128.021, 59.46] (p=0.458) |
| 2 | 0.310 | 10.166 (79.172) | 0.345 | 31.514 (170.872) | -21.348 [-80.216, 37.52] (p=0.467) |
| 3 | 0.639 | 4.761 (13.619) | 0.996 | 4.087 (7.497) | 0.674 [-2.259, 3.608] (p=0.65) |
| 4 | 1.560 | 15.946 (105.861) | 3.212 | 19.306 (93.554) | -3.361 [-35.804, 29.083] (p=0.837) |
| **5** | 2.971 | 8.515 (16.982) | 4.905 | 12.493 (27.809) | -3.978 [-12.724, 4.769] (p=0.365) |
| **6** | 4.248 | 14.622 (27.957) | 4.947 | 20.690 (41.400) | -6.068 [-19.829, 7.693] (p=0.379) |
| **7** | 6.795 | 19.451 (36.736) | 5.828 | 21.517 (52.349) | -2.067 [-17.89, 13.757] (p=0.795) |
| **8** | 8.276 | 18.669 (29.208) | 5.972 | 17.116 (28.221) | 1.553 [-7.321, 10.428] (p=0.728) |
| **9** | 10.452 | 21.630 (28.418) | 6.010 | 19.746 (28.660) | 1.884 [-7.399, 11.167] (p=0.687) |
| **10** | 11.346 | 21.729 (27.230) | 9.661 | 20.276 (26.774) | 1.453 [-7.444, 10.35] (p=0.745) |
| **11** | 8.845 | 20.584 (31.417) | 5.684 | 22.455 (40.704) | -1.871 [-14.866, 11.124] (p=0.774) |
| **12** | 7.947 | 18.386 (28.471) | 5.736 | 22.972 (39.378) | -4.586 [-17.357, 8.186] (p=0.475) |
| **13** | 3.544 | 11.815 (20.502) | 1.295 | 13.856 (23.400) | -2.041 [-9.843, 5.761] (p=0.603) |
| **14** | 2.607 | 10.200 (17.435) | 0.727 | 12.849 (24.050) | -2.649 [-11.476, 6.179] (p=0.548) |
| **Summary** | 3.391 | 15.058 (48.533) | 3.172 | 19.437 (72.080) | -4.379 [-10.508, 1.75] (p=0.161) |

**Supplementary Table 3 | CSF-Hb levels within 14 days of aneurysm rupture stratified by study site.**

The median, mean and the SD of CSF-Hb measurements (in µM) of the EVD and LD cohorts stratified by day post-SAH and study site.

#### *EVD cohort [median, mean (SD)]*

| Day after SAH | Site_A | Site_B | Site_C | Site_F | Site_G | Site_H | Site_E | Site_D |
|---|---|---|---|---|---|---|---|---|
| 1 | 2.98, 2.95 (1) | 3.18, 2.94 (1.08) | 3.93, 3.85 (0.77) | 2.8, 2.79 (1) | 2.7, 2.91 (1.16) | 3.15, 3.03 (1.05) | 2.96, 3.14 (0.43) | 3.01, 3.14 (1.04) |
| 2 | 2.95, 3.04 (0.89) | 2.55, 2.78 (0.89) | 2.84, 2.85 (0.56) | 3, 2.88 (1.25) | 3.51, 3.36 (0.58) | 3.3, 3.16 (1.44) | 2.98, 3.35 (1.38) | 3.41, 3.18 (1.03) |
| 3 | 3.03, 3.11 (1.08) | 2.52, 2.88 (1.45) | 2.72, 2.97 (0.96) | 2.82, 3 (1.11) | 3.36, 3.42 (0.62) | 3.19, 3.13 (1.08) | 2.75, 3.31 (1.76) | 2.97, 3.2 (1.21) |
| 4 | 2.94, 3.06 (1.07) | 2.95, 3.1 (1.05) | 3.07, 3.01 (0.96) | 3.05, 3.01 (1.14) | 3.03, 2.75 (1.27) | 3.08, 2.97 (1) | 3.29, 3.45 (1.28) | 3.15, 3.17 (1.19) |
| 5 | 3.14, 3.11 (1.01) | 2.67, 2.82 (0.85) | 2.76, 2.74 (0.84) | 2.82, 2.96 (1.1) | 2.83, 2.81 (0.81) | 2.96, 2.95 (0.77) | 3.06, 3.31 (1.49) | 3.18, 3.5 (1.18) |
| 6 | 3.02, 3.15 (1.07) | 3.09, 3.12 (1.08) | 2.78, 2.78 (0.86) | 2.96, 2.96 (1.03) | 2.92, 2.97 (0.93) | 2.29, 2.43 (1.07) | 3.42, 3.49 (1.21) | 2.94, 3.11 (1.11) |
| 7 | 2.86, 3.06 (1.27) | 3.19, 3.1 (0.9) | 3.42, 3.38 (1.47) | 2.68, 2.94 (1.03) | 3.05, 3.16 (1.07) | 3.3, 3.43 (1.17) | 3.08, 3.26 (1.87) | 2.9, 3.06 (1.42) |
| 8 | 2.68, 3.08 (1.4) | 3.09, 3.2 (1.24) | 2.74, 2.71 (1.22) | 3.34, 3.22 (1.13) | 2.63, 2.83 (1.04) | 3.14, 3.32 (1.49) | 3.93, 4.11 (1.55) | 2.74, 2.92 (1.48) |
| 9 | 3.57, 3.54 (1.44) | 2.9, 3.09 (0.86) | 3.48, 3.42 (0.8) | 3.15, 3.18 (1) | 3.03, 2.85 (0.91) | 3.41, 3.5 (1.54) | 3.19, 3.51 (1.56) | 2.58, 2.89 (1.46) |
| 10 | 3.12, 3.18 (1.49) | 3.01, 3.24 (0.77) | 2.91, 2.91 (0.81) | 3.15, 3.11 (1.13) | 3.06, 2.91 (1.08) | 3.65, 3.69 (1.39) | 3.42, 3.94 (1.8) | 3.2, 3.55 (1.33) |
| 11 | 3.53, 3.53 (1.61) | 3.08, 3.08 (0.78) | 2.6, 2.71 (1.17) | 2.54, 2.6 (0.86) | 3.02, 3.29 (0.93) | 3.17, 3.35 (1.41) | 2.89, 3.13 (1.12) | 3.07, 3.26 (1.1) |
| 12 | 3.34, 3.41 (1.51) | 3.23, 3.27 (0.91) | 3.39, 3.47 (0.9) | 2.93, 2.97 (1.04) | 3.56, 3.44 (1.06) | 3.66, 3.72 (1.27) | 3.15, 3.23 (1.55) | 3.19, 3.35 (1.46) |
| 13 | 3.29, 3.91 (1.77) | 3.01, 2.78 (0.95) | 3.42, 3.54 (1.2) | 2.94, 3.09 (1.22) | 3.14, 2.98 (1.04) | 2.78, 3.35 (1.73) | 2.98, 3.47 (1.9) | 3.39, 3.73 (1.48) |
| 14 | 3.25, 3.63 (1.79) | 3.99, 3.66 (0.8) | 3, 3.07 (0.97) | 2.63, 2.67 (0.98) | 3.37, 3.27 (1.15) | 3.23, 3.84 (2) | 3.25, 3.62 (1.62) | 3.31, 4.21 (2.14) |
| Summary | 3.06, 3.26 (1.35) | 3.01, 3.06 (0.97) | 2.99, 3.05 (1.03) | 2.94, 2.97 (1.08) | 3.07, 3.06 (0.99) | 3.18, 3.25 (1.32) | 3.19, 3.47 (1.52) | 3.07, 3.27 (1.33) |

### *LD cohort [median, mean (SD)]*

| Day after SAH | Site_A | Site_D | Site_C | Site_E | Site_F | Site_G | Site_H |
|---|---|---|---|---|---|---|---|
| 1 | 3.79,<br>4.51 (1.87) | 3.18,<br>3.89 (1.38) | NA,<br>NA (NA) | 3.38,<br>3.38 (0.76) | NA,<br>NA (NA) | 6.72,<br>6.72 (NA) | 5.92,<br>5.67 (1.87) |
| 2 | 3.69,<br>4.04 (1.31) | 3.45,<br>3.41 (1.55) | 2.33,<br>2.33 (NA) | 3.72,<br>4.32 (2.47) | 6.2,<br>6.2 (NA) | 6.59,<br>6.59 (NA) | 7.05,<br>6.01 (1.77) |
| 3 | 2.98,<br>3.72 (2.08) | 2.79,<br>3.61 (1.86) | 4.08,<br>4.08 (NA) | 3.19,<br>4.29 (2.17) | 6.74,<br>6.74 (NA) | 4.53,<br>4.53 (2.01) | 5.49,<br>4.92 (1.88) |
| 4 | 4.68,<br>4.48 (1.93) | 2.9,<br>3.2 (2.02) | NA,<br>NA (NA) | 4.92,<br>4.38 (1.86) | 5.62,<br>5.62 (NA) | 6.54,<br>6.55 (0.53) | 5.61,<br>5.23 (1.33) |
| 5 | 3.25,<br>3.84 (1.91) | 5.02,<br>4.75 (1.51) | 5.87,<br>5.87 (NA) | 5.43,<br>5.06 (1.73) | 8.38,<br>8.38 (NA) | 6.61,<br>6.6 (0.54) | 5.06,<br>4.83 (1.7) |
| 6 | 3.96,<br>4.44 (1.49) | 4,<br>4.48 (1.88) | 4.55,<br>4.55 (2.02) | 3.89,<br>4.58 (1.52) | 5.11,<br>5.11 (NA) | 4.86,<br>5.05 (0.45) | 4.94,<br>4.96 (2.1) |
| 7 | 3.83,<br>4.29 (2.13) | 4.32,<br>4.43 (2.15) | 5.63,<br>4.97 (2.63) | 4.2,<br>4.25 (2.4) | 4.96,<br>4.96 (NA) | 5.52,<br>6.01 (1.22) | 5.8,<br>5.69 (1.29) |
| 8 | 5.17,<br>4.38 (1.83) | 3.39,<br>3.66 (1.65) | 4.2,<br>4.2 (2.4) | 5.87,<br>5.2 (1.58) | 8.37,<br>8.37 (NA) | 6.32,<br>5.92 (1.11) | 6.02,<br>5.87 (1.3) |
| 9 | 4.97,<br>5.39 (1.76) | 2.07,<br>3.59 (2.45) | 5.78,<br>5.78 (2.66) | 4.79,<br>4.4 (2.34) | 6.56,<br>6.56 (NA) | 6.47,<br>6.47 (0.43) | 6.41,<br>6.45 (0.65) |
| 10 | 5.74,<br>5.07 (1.86) | 4.22,<br>4.45 (1.92) | 3.67,<br>4.43 (2.83) | 5.32,<br>5.61 (2) | 5.55,<br>5.55 (NA) | 6.76,<br>6.81 (0.13) | 6.46,<br>6.56 (1) |
| 11 | 5.72,<br>5.39 (1.63) | 3.8,<br>3.65 (1.74) | 4.99,<br>4.99 (2.05) | 3.31,<br>3.28 (1.12) | 5.07,<br>5.07 (NA) | 5.74,<br>5.74 (0.19) | 5.91,<br>6.12 (1.08) |
| 12 | 5.22,<br>5.35 (1.62) | 3.84,<br>4.42 (2.56) | 5.07,<br>5.07 (0.7) | 3.02,<br>3.56 (1.95) | 6.48,<br>6.48 (NA) | 6.71,<br>6.71 (NA) | 6.7,<br>6.76 (0.68) |
| 13 | 6.51,<br>6.23 (1.31) | 4.77,<br>5.06 (1.35) | 4.29,<br>4.29 (2.33) | 3.87,<br>4.44 (2.61) | 6.24,<br>6.24 (NA) | 5.88,<br>5.88 (NA) | 6.37,<br>6.18 (1.22) |
| 14 | 6.2,<br>6.04 (1.04) | 5.75,<br>5.38 (2.11) | 2.89,<br>2.89 (NA) | 3.25,<br>3.99 (2.35) | 4.66,<br>4.66 (NA) | 5.52,<br>5.52 (NA) | 5.79,<br>6.06 (1.24) |
| Summary | 5.02,<br>4.85 (1.84) | 4.08,<br>4.18 (1.88) | 4.33,<br>4.6 (1.84) | 3.9,<br>4.4 (1.96) | 6.2,<br>6.15 (1.19) | 6.16,<br>6.06 (0.93) | 6.07,<br>5.74 (1.48) |

**Supplementary Table 4 | Descriptive statistics on SAH-SBI-related outcomes stratified by study site.**

Incidence of subarachnoid hemorrhage-related secondary brain injury (SAH-SBI, i.e., composite outcome composed of aVSP and/or DCI and/or DIND), angiographic vasospasms (aVSP), delayed cerebral ischemia (DCI), delayed ischemic neurological deficits (DIND), triple H therapy (induction of hypertension and/or hypervolemia and/or hemodilution), spasmolysis and surgical decompression (hemicraniectomy or suboccipital craniectomy) in the full cohort, external ventricular drain (EVD) and lumbar drain (LD) cohorts. The absolute (n) and relative (%) incidence is given per patient and per assessment day (p, present; a, absent; m, missing).

| | Incidence | | | | | | | | | | | |
|---|---|---|---|---|---|---|---|---|---|---|---|---|
| | **Full cohort** | | | | **EVD cohort** | | | | **LD cohort** | | | |
| **Outcome Measure** | **Per patient** | **Per day** | | | **Per patient** | **Per day** | | | **Per patient** | **Per day** | | |
| | | **a** | **p** | **m** | | **a** | **p** | **m** | | **a** | **p** | **m** |
| **Site_A** | | | | | | | | | | | | |
| SAH-SBI | 71 (74.7) | 860 (66.1) | 191 (14.7) | 251 (19.3) | 51 (77.3) | 523 (58.4) | 148 (16.5) | 225 (25.1) | 16 (80.0) | 192 (68.6) | 51 (18.2) | 37 (13.2) |
| aVSP | 67 (70.5) | 183 (14.1) | 152 (11.7) | 967 (74.3) | 49 (74.2) | 140 (15.6) | 118 (13.2) | 638 (71.2) | 15 (75.0) | 36 (12.9) | 35 (12.5) | 209 (74.6) |
| DCI | 25 (26.3) | 297 (22.8) | 40 (3.1) | 965 (74.1) | 21 (31.8) | 224 (25.0) | 36 (4.0) | 636 (71.0) | 5 (25.0) | 66 (23.6) | 8 (2.9) | 206 (73.6) |
| DIND | 28 (29.5) | 821 (63.1) | 76 (5.8) | 405 (31.1) | 18 (27.3) | 475 (53.0) | 54 (6.0) | 367 (41.0) | 8 (40.0) | 197 (70.4) | 25 (8.9) | 58 (20.7) |
| Triple H | 60 (63.2) | 933 (71.7) | 358 (27.5) | 11 (0.8) | 44 (66.7) | 606 (67.6) | 286 (31.9) | 4 (0.4) | 13 (65.0) | 185 (66.1) | 94 (33.6) | 1 (0.4) |
| Spasmolysis | 22 (23.2) | 1256 (96.5) | 35 (2.7) | 11 (0.8) | 16 (24.2) | 865 (96.5) | 27 (3.0) | 4 (0.4) | 4 (20.0) | 274 (97.9) | 5 (1.8) | 1 (0.4) |
| Decompressi | 9 (9.5) | 1282 (98.5) | 9 (0.7) | 11 (0.8) | 7 (10.6) | 885 (98.8) | 7 (0.8) | 4 (0.4) | 1 (5.0) | 278 (99.3) | 1 (0.4) | 1 (0.4) |
| **Site_B** | | | | | | | | | | | | |
| SAH-SBI | 9 (47.4) | 146 (56.8) | 19 (7.4) | 92 (35.8) | 9 (47.4) | 146 (56.8) | 19 (7.4) | 92 (35.8) | | | | |
| aVSP | 5 (26.3) | 44 (17.1) | 10 (3.9) | 203 (79.0) | 5 (26.3) | 44 (17.1) | 10 (3.9) | 203 (79.0) | | | | |
| DCI | 4 (21.1) | 65 (25.3) | 8 (3.1) | 184 (71.6) | 4 (21.1) | 65 (25.3) | 8 (3.1) | 184 (71.6) | | | | |
| DIND | 2 (10.5) | 111 (43.2) | 2 (0.8) | 144 (56.0) | 2 (10.5) | 111 (43.2) | 2 (0.8) | 144 (56.0) | | | | |
| Triple H | 6 (31.6) | 216 (84.0) | 28 (10.9) | 13 (5.1) | 6 (31.6) | 216 (84.0) | 28 (10.9) | 13 (5.1) | | | | |
| Spasmolysis | 2 (10.5) | 242 (94.2) | 2 (0.8) | 13 (5.1) | 2 (10.5) | 242 (94.2) | 2 (0.8) | 13 (5.1) | | | | |
| Decompressi | 1 (5.3) | 243 (94.6) | 1 (0.4) | 13 (5.1) | 1 (5.3) | 243 (94.6) | 1 (0.4) | 13 (5.1) | | | | |
| **Site_C** | | | | | | | | | | | | |
| SAH-SBI | 21 (63.6) | 256 (55.7) | 59 (12.8) | 145 (31.5) | 18 (64.3) | 208 (53.3) | 54 (13.8) | 128 (32.8) | 0 (0.0) | 40 (95.2) | 0 (0.0) | 2 (4.8) |
| aVSP | 18 (54.5) | 114 (24.8) | 52 (11.3) | 294 (63.9) | 16 (57.1) | 98 (25.1) | 48 (12.3) | 244 (62.6) | 0 (0.0) | 10 (23.8) | 0 (0.0) | 32 (76.2) |
| DCI | 6 (18.2) | 160 (34.8) | 6 (1.3) | 294 (63.9) | 6 (21.4) | 140 (35.9) | 6 (1.5) | 244 (62.6) | 0 (0.0) | 10 (23.8) | 0 (0.0) | 32 (76.2) |
| DIND | 5 (15.2) | 193 (42.0) | 6 (1.3) | 261 (56.7) | 4 (14.3) | 148 (37.9) | 5 (1.3) | 237 (60.8) | 0 (0.0) | 39 (92.9) | 0 (0.0) | 3 (7.1) |
| Triple H | 1 (3.0) | 446 (97.0) | 3 (0.7) | 11 (2.4) | 1 (3.6) | 385 (98.7) | 3 (0.8) | 2 (0.5) | 0 (0.0) | 42 (100.0) | 0 (0.0) | 0 (0.0) |
| Spasmolysis | 7 (21.2) | 421 (91.5) | 28 (6.1) | 11 (2.4) | 6 (21.4) | 361 (92.6) | 27 (6.9) | 2 (0.5) | 0 (0.0) | 42 (100.0) | 0 (0.0) | 0 (0.0) |
| Decompressi | 5 (15.2) | 444 (96.5) | 5 (1.1) | 11 (2.4) | 5 (17.9) | 383 (98.2) | 5 (1.3) | 2 (0.5) | 0 (0.0) | 42 (100.0) | 0 (0.0) | 0 (0.0) |
| **Site_D** | | | | | | | | | | | | |
| SAH-SBI | 33 (53.2) | 663 (78.1) | 71 (8.4) | 115 (13.5) | 18 (58.1) | 315 (75.7) | 45 (10.8) | 56 (13.5) | 4 (40.0) | 112 (80.0) | 13 (9.3) | 15 (10.7) |
| aVSP | 28 (45.2) | 84 (9.9) | 49 (5.8) | 716 (84.3) | 16 (51.6) | 49 (11.8) | 28 (6.7) | 339 (81.5) | 4 (40.0) | 15 (10.7) | 7 (5.0) | 118 (84.3) |
| DCI | 10 (16.1) | 125 (14.7) | 17 (2.0) | 707 (83.3) | 7 (22.6) | 70 (16.8) | 12 (2.9) | 334 (80.3) | 2 (20.0) | 20 (14.3) | 3 (2.1) | 117 (83.6) |
| DIND | 16 (25.8) | 676 (79.6) | 36 (4.2) | 137 (16.1) | 11 (35.5) | 316 (76.0) | 30 (7.2) | 70 (16.8) | 3 (30.0) | 113 (80.7) | 9 (6.4) | 18 (12.9) |
| Triple H | 21 (33.9) | 718 (84.6) | 74 (8.7) | 57 (6.7) | 12 (38.7) | 355 (85.3) | 52 (12.5) | 9 (2.2) | 5 (50.0) | 118 (84.3) | 19 (13.6) | 3 (2.1) |
| Spasmolysis | 5 (8.1) | 779 (91.8) | 13 (1.5) | 57 (6.7) | 4 (12.9) | 395 (95.0) | 12 (2.9) | 9 (2.2) | 1 (10.0) | 134 (95.7) | 3 (2.1) | 3 (2.1) |
| Decompressi | 3 (4.8) | 789 (92.9) | 3 (0.4) | 57 (6.7) | 0 (0.0) | 407 (97.8) | 0 (0.0) | 9 (2.2) | 1 (10.0) | 136 (97.1) | 1 (0.7) | 3 (2.1) |
| **Site_E** | | | | | | | | | | | | |
| SAH-SBI | 2 (9.5) | 195 (68.2) | 5 (1.7) | 86 (30.1) | 1 (5.0) | 183 (67.3) | 3 (1.1) | 86 (31.6) | 1 (12.5) | 67 (59.8) | 2 (1.8) | 43 (38.4) |
| aVSP | 1 (4.8) | 34 (11.9) | 1 (0.3) | 251 (87.8) | 1 (5.0) | 33 (12.1) | 1 (0.4) | 238 (87.5) | 0 (0.0) | 15 (13.4) | 0 (0.0) | 97 (86.6) |
| DCI | 0 (0.0) | 36 (12.6) | 0 (0.0) | 250 (87.4) | 0 (0.0) | 35 (12.9) | 0 (0.0) | 237 (87.1) | 0 (0.0) | 15 (13.4) | 0 (0.0) | 97 (86.6) |
| DIND | 2 (9.5) | 167 (58.4) | 4 (1.4) | 115 (40.2) | 1 (5.0) | 156 (57.4) | 2 (0.7) | 114 (41.9) | 1 (12.5) | 51 (45.5) | 2 (1.8) | 59 (52.7) |
| Triple H | 0 (0.0) | 286 (100.0) | 0 (0.0) | 0 (0.0) | 0 (0.0) | 272 (100.0) | 0 (0.0) | 0 (0.0) | 0 (0.0) | 112 (100.0) | 0 (0.0) | 0 (0.0) |
| Spasmolysis | 1 (4.8) | 285 (99.7) | 1 (0.3) | 0 (0.0) | 1 (5.0) | 271 (99.6) | 1 (0.4) | 0 (0.0) | 1 (12.5) | 111 (99.1) | 1 (0.9) | 0 (0.0) |
| Decompressi | 5 (23.8) | 280 (97.9) | 6 (2.1) | 0 (0.0) | 5 (25.0) | 266 (97.8) | 6 (2.2) | 0 (0.0) | 1 (12.5) | 111 (99.1) | 1 (0.9) | 0 (0.0) |
| **Site_F** | | | | | | | | | | | | |
| SAH-SBI | 36 (59.0) | 630 (75.6) | 100 (12.0) | 103 (12.4) | 32 (64.0) | 489 (72.0) | 92 (13.5) | 98 (14.4) | 0 (0.0) | 14 (100.0) | 0 (0.0) | 0 (0.0) |
| aVSP | 25 (41.0) | 187 (22.4) | 52 (6.2) | 594 (71.3) | 22 (44.0) | 164 (24.2) | 45 (6.6) | 470 (69.2) | 0 (0.0) | 1 (7.1) | 0 (0.0) | 13 (92.9) |
| DCI | 23 (37.7) | 232 (27.9) | 49 (5.9) | 552 (66.3) | 20 (40.0) | 203 (29.9) | 46 (6.8) | 430 (63.3) | 0 (0.0) | 3 (21.4) | 0 (0.0) | 11 (78.6) |
| DIND | 26 (42.6) | 599 (71.9) | 59 (7.1) | 175 (21.0) | 23 (46.0) | 456 (67.2) | 53 (7.8) | 170 (25.0) | 0 (0.0) | 14 (100.0) | 0 (0.0) | 0 (0.0) |
| Triple H | 50 (82.0) | 340 (40.8) | 473 (56.8) | 20 (2.4) | 43 (86.0) | 246 (36.2) | 417 (61.4) | 16 (2.4) | 1 (100.0) | 10 (71.4) | 4 (28.6) | 0 (0.0) |
| Spasmolysis | 24 (39.3) | 749 (89.9) | 64 (7.7) | 20 (2.4) | 21 (42.0) | 611 (90.0) | 52 (7.7) | 16 (2.4) | 0 (0.0) | 14 (100.0) | 0 (0.0) | 0 (0.0) |
| Decompressi | 11 (18.0) | 801 (96.2) | 12 (1.4) | 20 (2.4) | 11 (22.0) | 651 (95.9) | 12 (1.8) | 16 (2.4) | 0 (0.0) | 14 (100.0) | 0 (0.0) | 0 (0.0) |
| **Site_G** | | | | | | | | | | | | |
| SAH-SBI | 11 (61.1) | 172 (68.3) | 24 (9.5) | 56 (22.2) | 9 (60.0) | 142 (67.6) | 17 (8.1) | 51 (24.3) | 2 (66.7) | 30 (71.4) | 7 (16.7) | 5 (11.9) |
| aVSP | 8 (44.4) | 43 (17.1) | 18 (7.1) | 191 (75.8) | 6 (40.0) | 38 (18.1) | 11 (5.2) | 161 (76.7) | 2 (66.7) | 5 (11.9) | 7 (16.7) | 30 (71.4) |
| DCI | 7 (38.9) | 63 (25.0) | 9 (3.6) | 180 (71.4) | 6 (40.0) | 53 (25.2) | 6 (2.9) | 151 (71.9) | 1 (33.3) | 10 (23.8) | 3 (7.1) | 29 (69.0) |
| DIND | 5 (27.8) | 148 (58.7) | 5 (2.0) | 99 (39.3) | 5 (33.3) | 114 (54.3) | 5 (2.4) | 91 (43.3) | 0 (0.0) | 34 (81.0) | 0 (0.0) | 8 (19.0) |
| Triple H | 0 (0.0) | 247 (98.0) | 0 (0.0) | 5 (2.0) | 0 (0.0) | 208 (99.0) | 0 (0.0) | 2 (1.0) | 0 (0.0) | 39 (92.9) | 0 (0.0) | 3 (7.1) |
| Spasmolysis | 6 (33.3) | 224 (88.9) | 23 (9.1) | 5 (2.0) | 4 (26.7) | 197 (93.8) | 11 (5.2) | 2 (1.0) | 2 (66.7) | 27 (64.3) | 12 (28.6) | 3 (7.1) |
| Decompressi | 2 (11.1) | 245 (97.2) | 2 (0.8) | 5 (2.0) | 2 (13.3) | 206 (98.1) | 2 (1.0) | 2 (1.0) | 0 (0.0) | 39 (92.9) | 0 (0.0) | 3 (7.1) |
| **Site_H** | | | | | | | | | | | | |
| SAH-SBI | 26 (45.6) | 646 (81.0) | 65 (8.1) | 87 (10.9) | 16 (51.6) | 352 (81.1) | 38 (8.8) | 44 (10.1) | 12 (57.1) | 249 (84.7) | 29 (9.9) | 16 (5.4) |
| aVSP | 17 (29.8) | 72 (9.0) | 33 (4.1) | 693 (86.8) | 9 (29.0) | 46 (10.6) | 19 (4.4) | 369 (85.0) | 9 (42.9) | 20 (6.8) | 15 (5.1) | 259 (88.1) |
| DCI | 8 (14.0) | 127 (15.9) | 13 (1.6) | 658 (82.5) | 6 (19.4) | 78 (18.0) | 10 (2.3) | 346 (79.7) | 2 (9.5) | 49 (16.7) | 3 (1.0) | 242 (82.3) |
| DIND | 18 (31.6) | 640 (80.2) | 40 (5.0) | 118 (14.8) | 10 (32.3) | 341 (78.6) | 20 (4.6) | 73 (16.8) | 10 (47.6) | 254 (86.4) | 22 (7.5) | 18 (6.1) |
| Triple H | 17 (29.8) | 663 (83.1) | 75 (9.4) | 60 (7.5) | 13 (41.9) | 358 (82.5) | 53 (12.2) | 23 (5.3) | 6 (28.6) | 254 (86.4) | 25 (8.5) | 15 (5.1) |
| Spasmolysis | 8 (14.0) | 721 (90.4) | 17 (2.1) | 60 (7.5) | 5 (16.1) | 397 (91.5) | 14 (3.2) | 23 (5.3) | 4 (19.0) | 275 (93.5) | 4 (1.4) | 15 (5.1) |
| Decompressi | 7 (12.3) | 731 (91.6) | 7 (0.9) | 60 (7.5) | 5 (16.1) | 406 (93.5) | 5 (1.2) | 23 (5.3) | 2 (9.5) | 277 (94.2) | 2 (0.7) | 15 (5.1) |

**Supplementary Table 5 | Descriptive statistics on outcomes at three-month follow-up visit.**

Incidence (n, %) of chronic hydrocephalus (defined as dependency on a ventriculoperitoneal shunt) and functional status (GOSE, Glascow Outcome Scale-Extended; mRS, modified Rankin Scale) at three-months follow-up visits.

| Measure | Overall | Site_A | Site_B | Site_C | Site_D | Site_E | Site_F | Site_G | Site_H |
|---|---|---|---|---|---|---|---|---|---|
| **Full cohort** | | | | | | | | | |
| **N (cohort size)** | 366 | 95 | 19 | 33 | 62 | 21 | 61 | 18 | 57 |
| **Chronic hydrocephalus at 12 weeks** | 90 (24.6%) | 34 (35.8%) | 4 (21.1%) | 11 (33.3%) | 17 (27.4%) | 6 (28.6%) | 10 (16.4%) | 2 (11.1%) | 6 (10.5%) |
| **GOSE** | | | | | | | | | |
| 1 | 47 (12.8%) | 10 (10.5%) | 3 (15.8%) | 4 (12.1%) | 7 (11.3%) | 5 (23.8%) | 8 (13.1%) | 3 (16.7%) | 7 (12.3%) |
| 2 | 8 (2.2%) | 2 (2.1%) | 0 (0.0%) | 1 (3.0%) | 1 (1.6%) | 2 (9.5%) | 0 (0.0%) | 1 (5.6%) | 1 (1.8%) |
| 3 | 52 (14.2%) | 13 (13.7%) | 0 (0.0%) | 8 (24.2%) | 5 (8.1%) | 2 (9.5%) | 14 (23.0%) | 7 (38.9%) | 3 (5.3%) |
| 4 | 24 (6.6%) | 7 (7.4%) | 3 (15.8%) | 2 (6.1%) | 2 (3.2%) | 1 (4.8%) | 7 (11.5%) | 1 (5.6%) | 1 (1.8%) |
| 5 | 13 (3.6%) | 1 (1.1%) | 0 (0.0%) | 0 (0.0%) | 2 (3.2%) | 1 (4.8%) | 7 (11.5%) | 0 (0.0%) | 2 (3.5%) |
| 6 | 32 (8.7%) | 10 (10.5%) | 3 (15.8%) | 2 (6.1%) | 8 (12.9%) | 2 (9.5%) | 5 (8.2%) | 0 (0.0%) | 2 (3.5%) |
| 7 | 109 (29.8%) | 31 (32.6%) | 4 (21.1%) | 10 (30.3%) | 26 (41.9%) | 2 (9.5%) | 14 (23.0%) | 2 (11.1%) | 20 (35.1%) |
| 8 | 80 (21.9%) | 20 (21.1%) | 6 (31.6%) | 6 (18.2%) | 11 (17.7%) | 6 (28.6%) | 6 (9.8%) | 4 (22.2%) | 21 (36.8%) |
| **mRS** | | | | | | | | | |
| 0 | 56 (15.3%) | 16 (16.8%) | 7 (36.8%) | 5 (15.2%) | 7 (11.3%) | 2 (9.5%) | 3 (4.9%) | 6 (33.3%) | 10 (17.5%) |
| 1 | 103 (28.1%) | 25 (26.3%) | 5 (26.3%) | 7 (21.2%) | 24 (38.7%) | 6 (28.6%) | 12 (19.7%) | 0 (0.0%) | 24 (42.1%) |
| 2 | 62 (16.9%) | 21 (22.1%) | 2 (10.5%) | 5 (15.2%) | 11 (17.7%) | 1 (4.8%) | 14 (23.0%) | 1 (5.6%) | 7 (12.3%) |
| 3 | 36 (9.8%) | 8 (8.4%) | 2 (10.5%) | 0 (0.0%) | 8 (12.9%) | 2 (9.5%) | 11 (18.0%) | 2 (11.1%) | 3 (5.3%) |
| 4 | 39 (10.7%) | 10 (10.5%) | 0 (0.0%) | 7 (21.2%) | 3 (4.8%) | 3 (14.3%) | 9 (14.8%) | 4 (22.2%) | 3 (5.3%) |
| 5 | 22 (6.0%) | 4 (4.2%) | 0 (0.0%) | 5 (15.2%) | 2 (3.2%) | 2 (9.5%) | 4 (6.6%) | 2 (11.1%) | 3 (5.3%) |
| 6 | 47 (12.8%) | 10 (10.5%) | 3 (15.8%) | 4 (12.1%) | 7 (11.3%) | 5 (23.8%) | 8 (13.1%) | 3 (16.7%) | 7 (12.3%) |
| **Lost to follow-up at 12 weeks** | 1 (0.3%) | 1 (1.1%) | 0 (0.0%) | 0 (0.0%) | 0 (0.0%) | 0 (0.0%) | 0 (0.0%) | 0 (0.0%) | 0 (0.0%) |
| **EVD cohort** | | | | | | | | | |
| **N (cohort size)** | 260 | 66 | 19 | 28 | 31 | 20 | 50 | 15 | 31 |
| **Chronic hydrocephalus at 12 weeks** | 84 (32.3%) | 32 (48.5%) | 4 (21.1%) | 11 (39.3%) | 15 (48.4%) | 6 (30.0%) | 9 (18.0%) | 2 (13.3%) | 5 (16.1%) |
| **GOSE** | | | | | | | | | |
| 1 | 43 (16.5%) | 9 (13.6%) | 3 (15.8%) | 4 (14.3%) | 6 (19.4%) | 5 (25.0%) | 8 (16.0%) | 2 (13.3%) | 6 (19.4%) |
| 2 | 8 (3.1%) | 2 (3.0%) | 0 (0.0%) | 1 (3.6%) | 1 (3.2%) | 2 (10.0%) | 0 (0.0%) | 1 (6.7%) | 1 (3.2%) |
| 3 | 48 (18.5%) | 12 (18.2%) | 0 (0.0%) | 8 (28.6%) | 3 (9.7%) | 2 (10.0%) | 14 (28.0%) | 6 (40.0%) | 3 (9.7%) |
| 4 | 20 (7.7%) | 7 (10.6%) | 3 (15.8%) | 1 (3.6%) | 1 (3.2%) | 1 (5.0%) | 6 (12.0%) | 1 (6.7%) | 0 (0.0%) |
| 5 | 8 (3.1%) | 1 (1.5%) | 0 (0.0%) | 0 (0.0%) | 1 (3.2%) | 1 (5.0%) | 4 (8.0%) | 0 (0.0%) | 1 (3.2%) |
| 6 | 20 (7.7%) | 3 (4.5%) | 3 (15.8%) | 2 (7.1%) | 6 (19.4%) | 1 (5.0%) | 4 (8.0%) | 0 (0.0%) | 1 (3.2%) |
| 7 | 66 (25.4%) | 21 (31.8%) | 4 (21.1%) | 7 (25.0%) | 9 (29.0%) | 2 (10.0%) | 11 (22.0%) | 2 (13.3%) | 10 (32.3%) |
| 8 | 46 (17.7%) | 10 (15.2%) | 6 (31.6%) | 5 (17.9%) | 4 (12.9%) | 6 (30.0%) | 3 (6.0%) | 3 (20.0%) | 9 (29.0%) |
| **mRS** | | | | | | | | | |
| 0 | 34 (13.1%) | 9 (13.6%) | 7 (36.8%) | 4 (14.3%) | 3 (9.7%) | 2 (10.0%) | 1 (2.0%) | 5 (33.3%) | 3 (9.7%) |
| 1 | 56 (21.5%) | 16 (24.2%) | 5 (26.3%) | 4 (14.3%) | 6 (19.4%) | 6 (30.0%) | 7 (14.0%) | 0 (0.0%) | 12 (38.7%) |
| 2 | 48 (18.5%) | 14 (21.2%) | 2 (10.5%) | 5 (17.9%) | 8 (25.8%) | 1 (5.0%) | 14 (28.0%) | 1 (6.7%) | 3 (9.7%) |
| 3 | 21 (8.1%) | 3 (4.5%) | 2 (10.5%) | 0 (0.0%) | 4 (12.9%) | 1 (5.0%) | 7 (14.0%) | 2 (13.3%) | 2 (6.5%) |
| 4 | 36 (13.8%) | 10 (15.2%) | 0 (0.0%) | 6 (21.4%) | 3 (9.7%) | 3 (15.0%) | 9 (18.0%) | 3 (20.0%) | 2 (6.5%) |
| 5 | 21 (8.1%) | 4 (6.1%) | 0 (0.0%) | 5 (17.9%) | 1 (3.2%) | 2 (10.0%) | 4 (8.0%) | 2 (13.3%) | 3 (9.7%) |
| 6 | 43 (16.5%) | 9 (13.6%) | 3 (15.8%) | 4 (14.3%) | 6 (19.4%) | 5 (25.0%) | 8 (16.0%) | 2 (13.3%) | 6 (19.4%) |
| **Lost to follow-up at 12 weeks** | 1 (0.4%) | 1 (1.5%) | 0 (0.0%) | 0 (0.0%) | 0 (0.0%) | 0 (0.0%) | 0 (0.0%) | 0 (0.0%) | 0 (0.0%) |
| **LD cohort** | | | | | | | | | |
| **N (cohort size)** | 66 | 20 | 0 | 3 | 10 | 8 | 1 | 3 | 21 |
| **Chronic hydrocephalus at 12 weeks** | 19 (28.8%) | 10 (50.0%) | 0 (0%) | 1 (33.3%) | 5 (50.0%) | 1 (12.5%) | 1 (100.0%) | 0 (0.0%) | 1 (4.8%) |
| **GOSE** | | | | | | | | | |
| 1 | 5 (7.6%) | 0 (0.0%) | 0 (0%) | 0 (0.0%) | 1 (10.0%) | 2 (25.0%) | 0 (0.0%) | 1 (33.3%) | 1 (4.8%) |
| 2 | 1 (1.5%) | 0 (0.0%) | 0 (0%) | 0 (0.0%) | 0 (0.0%) | 1 (12.5%) | 0 (0.0%) | 0 (0.0%) | 0 (0.0%) |
| 3 | 6 (9.1%) | 3 (15.0%) | 0 (0%) | 0 (0.0%) | 1 (10.0%) | 1 (12.5%) | 0 (0.0%) | 1 (33.3%) | 0 (0.0%) |
| 4 | 4 (6.1%) | 2 (10.0%) | 0 (0%) | 0 (0.0%) | 0 (0.0%) | 0 (0.0%) | 1 (100.0%) | 0 (0.0%) | 1 (4.8%) |
| 5 | 2 (3.0%) | 0 (0.0%) | 0 (0%) | 0 (0.0%) | 1 (10.0%) | 0 (0.0%) | 0 (0.0%) | 0 (0.0%) | 1 (4.8%) |
| 6 | 4 (6.1%) | 1 (5.0%) | 0 (0%) | 0 (0.0%) | 1 (10.0%) | 1 (12.5%) | 0 (0.0%) | 0 (0.0%) | 1 (4.8%) |
| 7 | 26 (39.4%) | 10 (50.0%) | 0 (0%) | 2 (66.7%) | 4 (40.0%) | 1 (12.5%) | 0 (0.0%) | 0 (0.0%) | 9 (42.9%) |
| 8 | 18 (27.3%) | 4 (20.0%) | 0 (0%) | 1 (33.3%) | 2 (20.0%) | 2 (25.0%) | 0 (0.0%) | 1 (33.3%) | 8 (38.1%) |
| **mRS** | | | | | | | | | |
| 0 | 13 (19.7%) | 6 (30.0%) | 0 (0%) | 1 (33.3%) | 1 (10.0%) | 0 (0.0%) | 0 (0.0%) | 1 (33.3%) | 4 (19.0%) |
| 1 | 23 (34.8%) | 5 (25.0%) | 0 (0%) | 2 (66.7%) | 4 (40.0%) | 3 (37.5%) | 0 (0.0%) | 0 (0.0%) | 9 (42.9%) |
| 2 | 12 (18.2%) | 5 (25.0%) | 0 (0%) | 0 (0.0%) | 2 (20.0%) | 0 (0.0%) | 0 (0.0%) | 0 (0.0%) | 5 (23.8%) |
| 3 | 7 (10.6%) | 3 (15.0%) | 0 (0%) | 0 (0.0%) | 1 (10.0%) | 1 (12.5%) | 1 (100.0%) | 0 (0.0%) | 1 (4.8%) |
| 4 | 5 (7.6%) | 1 (5.0%) | 0 (0%) | 0 (0.0%) | 1 (10.0%) | 1 (12.5%) | 0 (0.0%) | 1 (33.3%) | 1 (4.8%) |
| 5 | 1 (1.5%) | 0 (0.0%) | 0 (0%) | 0 (0.0%) | 0 (0.0%) | 1 (12.5%) | 0 (0.0%) | 0 (0.0%) | 0 (0.0%) |
| 6 | 5 (7.6%) | 0 (0.0%) | 0 (0%) | 0 (0.0%) | 1 (10.0%) | 2 (25.0%) | 0 (0.0%) | 1 (33.3%) | 1 (4.8%) |
| **Lost to follow-up at 12 weeks** | 0 (0.0%) | 0 (0.0%) | 0 (0%) | 0 (0.0%) | 0 (0.0%) | 0 (0.0%) | 0 (0.0%) | 0 (0.0%) | 0 (0.0%) |

**Supplementary Table 6 | Daily CSF-metHb levels within 14 days of aneurysm rupture.**

The median, mean and the SD of CSF-metHb measurements (in µM) within the total (multicenter) EVD cohort stratified by day post-SAH.

| *Day post SAH* | *EVD (median)* | *EVD (mean [SD])* |
|---|---|---|
| 1 | 0.1 | 0.165 (0.459) |
| 2 | 0.1 | 0.233 (0.907) |
| 3 | 0.1 | 0.445 (2.025) |
| 4 | 0.1 | 0.649 (2.472) |
| 5 | 0.1 | 0.879 (1.934) |
| 6 | 0.123 | 0.934 (1.708) |
| 7 | 0.353 | 1.567 (3.540) |
| 8 | 0.61 | 1.801 (3.190) |
| 9 | 0.974 | 2.488 (4.598) |
| 10 | 1.40 | 3.053 (4.673) |
| 11 | 1.35 | 3.085 (5.145) |
| 12 | 1.41 | 3.126 (4.560) |
| 13 | 1.00 | 2.443 (4.303) |
| 14 | 0.987 | 2.219 (3.237) |

**Supplementary Table 7 | Descriptive statistics on safety assessments.**

Incidence (n, %) of cerebrospinal fluid (CSF) infection (clinical symptoms and signs of CSF infection and positive CSF culture), colonization (two positive CSF cultures without clinical symptoms or signs), contamination (1 positive CSF culture with consecutive culture being negative) or wound revision due to surgical site infection at the EVD/LD skin entrance.

| Safety endpoints | Overall | Site_A | Site_D | Site_C | Site_F | Site_E | Site_B | Site_G | Site_H |
|---|---|---|---|---|---|---|---|---|---|
| **Full cohort** | | | | | | | | | |
| n | 366 | 95 | 62 | 33 | 61 | 21 | 19 | 18 | 57 |
| CSF infection | 8 (2.2%) | 0 (0%) | 4 (6.5%) | 0 (0%) | 0 (0%) | 0 (0%) | 1 (5.3%) | 0 (0%) | 3 (5.3%) |
| Colonisation | 2 (0.5%) | 1 (1.1%) | 0 (0%) | 0 (0%) | 0 (0%) | 0 (0%) | 1 (5.3%) | 0 (0%) | 0 (0%) |
| Contamination | 0 (0%) | 0 (0%) | 0 (0%) | 0 (0%) | 0 (0%) | 0 (0%) | 0 (0%) | 0 (0%) | 0 (0%) |
| Wound revision | 7 (1.9%) | 3 (3.2%) | 1 (1.6%) | 0 (0%) | 2 (3.3%) | 0 (0%) | 0 (0%) | 0 (0%) | 1 (1.8%) |
| **EVD cohort** | | | | | | | | | |
| n | 260 | 66 | 31 | 28 | 50 | 20 | 19 | 15 | 31 |
| CSF infection | 5 (1.9%) | 0 (0%) | 3 (9.7%) | 0 (0%) | 0 (0%) | 0 (0%) | 1 (5.3%) | 0 (0%) | 1 (3.2%) |
| Colonisation | 1 (0.4%) | 0 (0%) | 0 (0%) | 0 (0%) | 0 (0%) | 0 (0%) | 1 (5.3%) | 0 (0%) | 0 (0%) |
| Contamination | 0 (0%) | 0 (0%) | 0 (0%) | 0 (0%) | 0 (0%) | 0 (0%) | 0 (0%) | 0 (0%) | 0 (0%) |
| Wound revision | 5 (1.9%) | 2 (3%) | 0 (0%) | 0 (0%) | 2 (4%) | 0 (0%) | 0 (0%) | 0 (0%) | 1 (3.2%) |
| **LD cohort** | | | | | | | | | |
| n | 66 | 20 | 10 | 3 | 1 | 8 | 0 | 3 | 21 |
| CSF infection | 4 (6.1%) | 0 (0%) | 2 (20%) | 0 (0%) | 0 (0%) | 0 (0%) | 0 (0%) | 0 (0%) | 2 (9.5%) |
| Colonisation | 1 (1.5%) | 1 (5%) | 0 (0%) | 0 (0%) | 0 (0%) | 0 (0%) | 0 (0%) | 0 (0%) | 0 (0%) |
| Contamination | 0 (0%) | 0 (0%) | 0 (0%) | 0 (0%) | 0 (0%) | 0 (0%) | 0 (0%) | 0 (0%) | 0 (0%) |
| Wound revision | 2 (3%) | 2 (10%) | 0 (0%) | 0 (0%) | 0 (0%) | 0 (0%) | 0 (0%) | 0 (0%) | 0 (0%) |

### Supplementary Figures

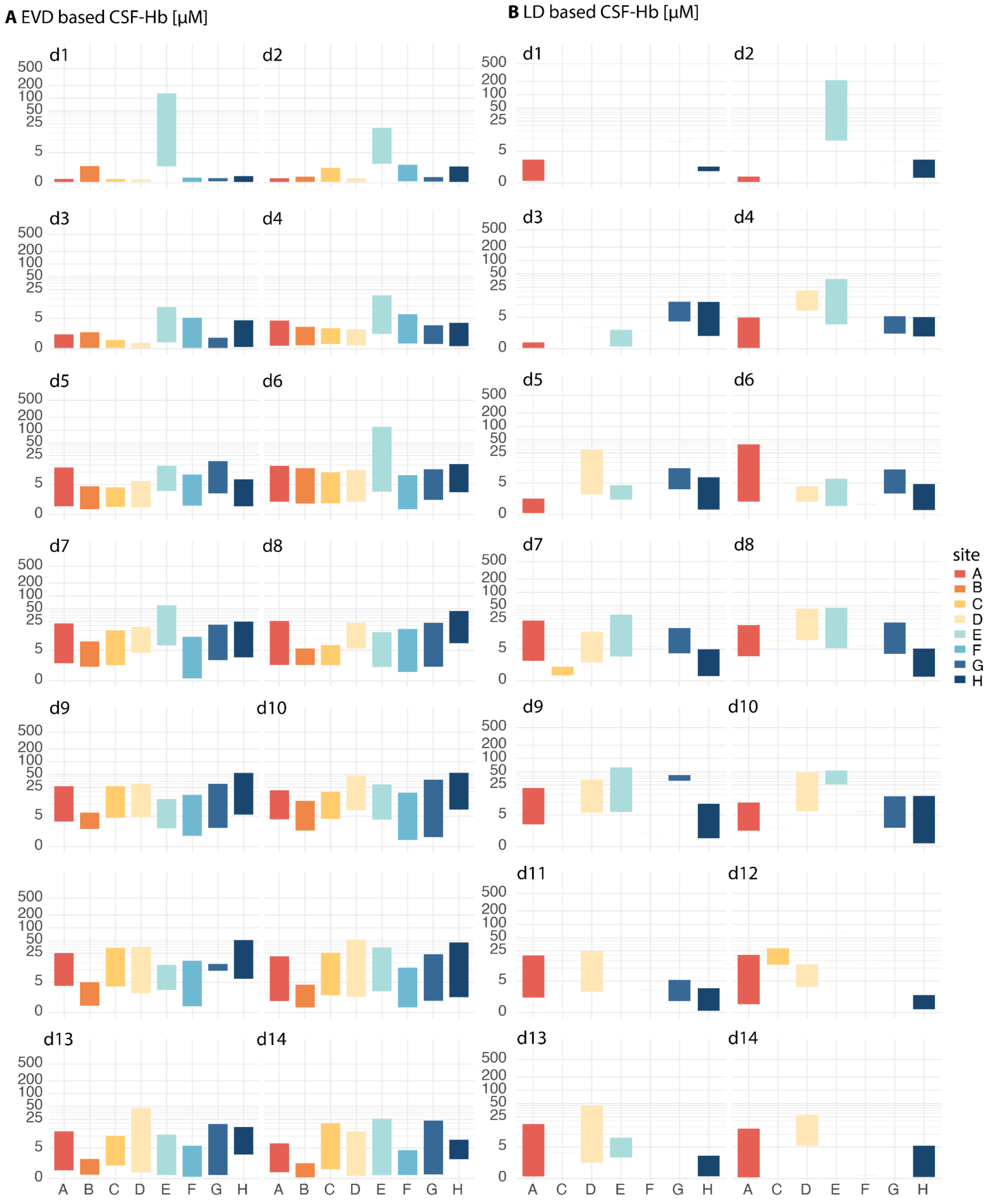


**Supplementary Figure 1 | EVD- and LD-derived CSF-Hb on each day post-SAH stratified by study site.**

(**A**) Daily EVD-derived CSF-Hb and (**B**) LD-derived CSF-Hb concentrations stratified by study site and day after haemorrhage reported as IQR. Wide inter-site variability was observed.

*Abbreviations: CSF-Hb, cerebrospinal fluid oxyhaemoglobin; EVD, external ventricular drain; LD, lumbar drain; aSAH, aneurysmal subarachnoid haemorrhage.*

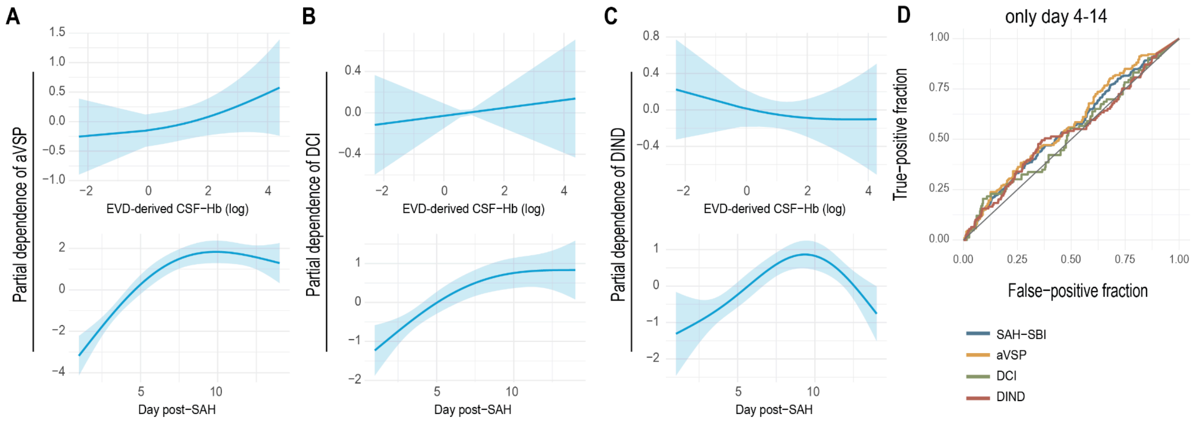


**Supplementary Figure 2 | Association of EVD-derived CSF-Hb with secondary brain injury**

(**A–C**) Partial-dependence plots from GAMs showing no significant associations between EVD-based CSF-Hb and aVSP, DCI, or DIND. (**D**) Receiver-operating characteristic curves during the high-risk phase (days 4–14) demonstrating poor discrimination (AUC < 0.6 for all).

*Abbreviations: CSF-Hb, cerebrospinal fluid oxyhaemoglobin; EVD, external ventricular drain; aVSP, angiographic vasospasm; DCI, delayed cerebral ischaemia; DIND, delayed ischaemic neurological deficit; GAM, generalised additive model; AUC, area under the curve.*

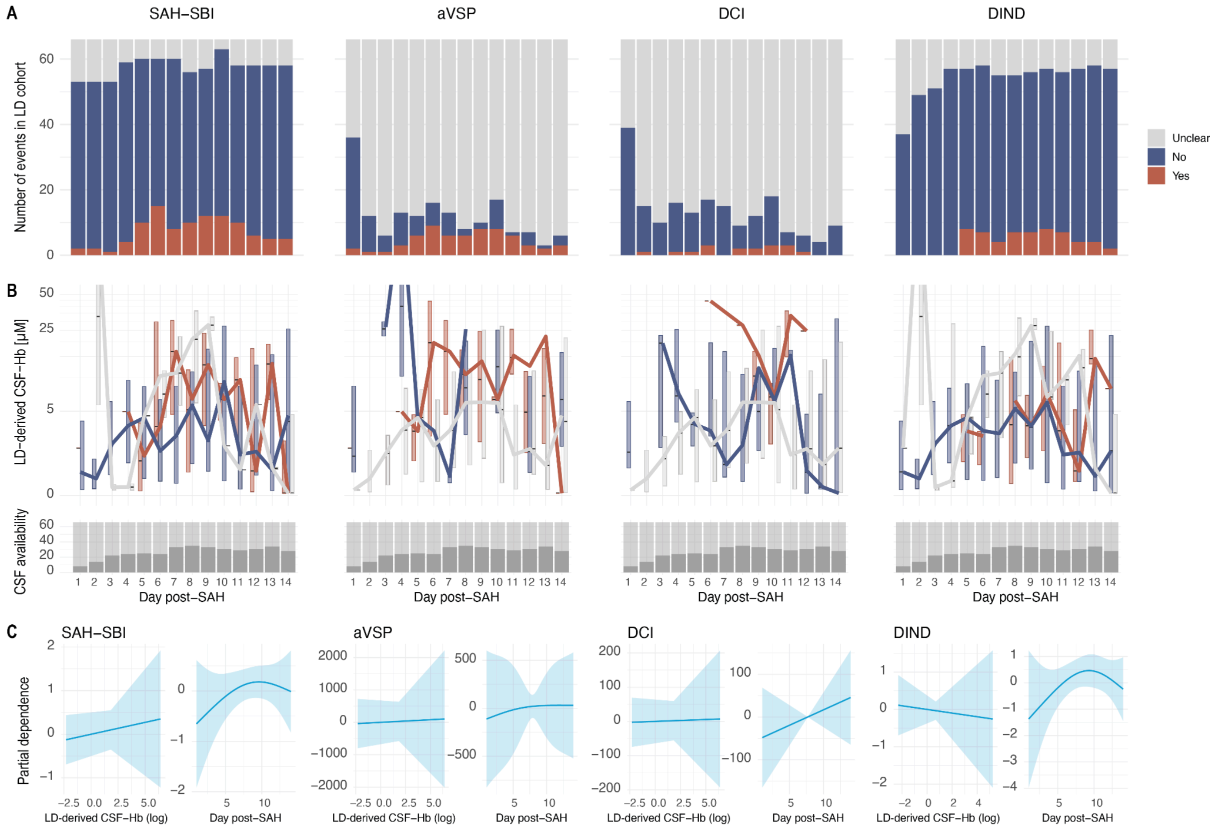


**Supplementary Figure 3 | LD-derived CSF-Hb and secondary brain injury.**

(**A**) Daily incidence of SAH-SBI, aVSP, DCI, and DIND in the LD cohort. (**B**) Temporal CSF-Hb trajectories stratified by clinical events (top). Availability of CSF samples (bottom). (**C**) Partial-dependence plots from GAMs showing no significant associations between LD-derived CSF-Hb and any outcome.

*Abbreviations: LD, lumbar drain; CSF-Hb, cerebrospinal fluid oxyhaemoglobin; SAH-SBI, subarachnoid haemorrhage–related secondary brain injury; aVSP, angiographic vasospasm; DCI, delayed cerebral ischaemia; DIND, delayed ischaemic neurological deficit.*

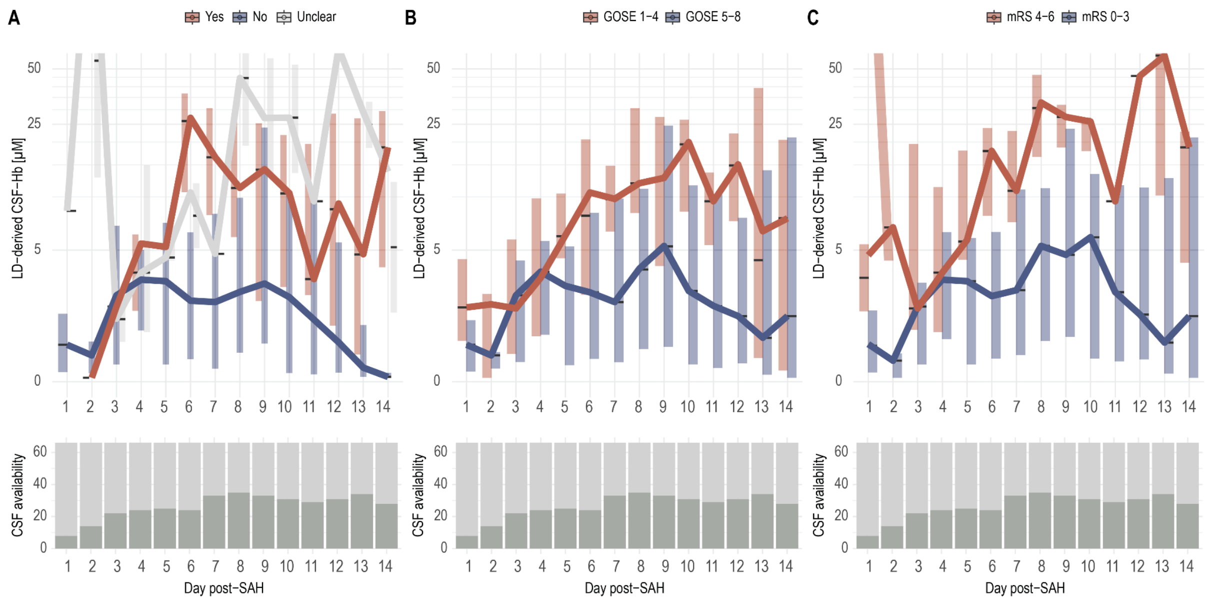


**Supplementary Figure 4 | LD-derived CSF-Hb and 3-month follow-up.**

(**A**) Temporal course of LD-derived CSF-Hb stratified by chronic hydrocephalus. (**B**) CSF-Hb levels by GOSE category. (**C**) Corresponding stratification by mRS at 3 months. Higher acute-phase CSF-Hb was observed in patients with chronic hydrocephalus or poor outcomes. Bottom panels depict the corresponding availability of CSF samples.

*Abbreviations: LD, lumbar drain; CSF-Hb, cerebrospinal fluid oxyhaemoglobin; GOSE, Glasgow Outcome Scale-Extended; mRS, modified Rankin Scale; cHCP, chronic hydrocephalus.*
